# Distribution of tick-borne microorganisms in human-biting ticks in France collected through a Citizen-science program

**DOI:** 10.1101/2025.09.22.25336155

**Authors:** Jonas Durand, Thierno Bah Madiou, Isabelle Lebert, Clémence Galon, Irene Carravieri, Sébastien Masseglia, Jean-Marc Armand, Julien Marchand, Cyril Galley, Karine Chalvet-Monfray, Muriel Vayssier-Taussat, Gwenaël Vourc’h, Annick Brun-Jacob, Sara Moutailler, Xavier Bailly, Pascale Frey-Klett

## Abstract

Ticks occupy diverse habitats, increasing the risk of human exposure. Assessing the public health threat posed by ticks requires rigorous monitoring of their distribution and of the prevalence of tick-borne pathogens. In France since 2017, the citizen science program CiTIQUE monitors human tick bites through multiple complementary approaches. Citizens can report bites and submit biting ticks to a national tick bank for research and surveillance. This study aimed to investigate human exposure to tick-borne microorganisms including pathogens across France, using ticks submitted through the CiTIQUE program. In total, 2,009 ticks were selected from the CiTIQUE tick bank, identified, and screened for microorganisms using a real-time microfluidic PCR method. Most bites involved *Ixodes ricinus* nymphs except in Mediterranean regions where *Dermacentor* and *Rhipicephalus* ticks were more common. Twenty-six microorganisms were detected, eighteen of which are potentially pathogenic to humans. These pathogens were widely distributed across the country. *Borrelia spp.* were the most frequently detected pathogens with spatial variation among regions. *Anaplasma phagocytophilum* infection rates varied from region to region. *Neoehrlichia mikurensis* was found in seven out of twelve French regions. *Rickettsia* species diversity was highest in the southeast, associated with a greater diversity of vectors. Five percent of ticks were infected with more than one pathogen. Although spatial heterogeneity was observed, no region was free of infected ticks. This study demonstrates the power of citizen science for nationwide surveillance of tick-borne pathogens, providing a large-scale overview of pathogen diversity and distribution across France from crowdsourced tick data.

## Introduction

Ticks of the family *Ixodidae*, commonly known as hard ticks, are major vectors of infectious diseases worldwide. They pose a substantial public health risk, for instance in temperate regions such as western Europe (Pustijanac et al., 2024). These ectoparasites can transmit a diverse range of pathogens, including bacteria, viruses, and protozoa (Guglielmone and Robbins, 2018). While a species can be recognized as pathogenic, there are usually important variation in its pathogenicity between different strains of the same species.

In Europe, Lyme borreliosis is the most prevalent tick-borne disease. It is caused by bacteria of the *Borrelia burgdorferi* sensu lato (Bbsl) complex, primarily transmitted by *Ixodes ricinus,* the most widespread tick species in the area (Kilpatrick et al., 2017). These bacteria can infect multiple organs, leading to dermatological, neurological, articular, or cardiac manifestations in humans. Of the 21 genospecies existing worldwide, seven - *Borrelia afzelii*, *Borrelia garinii*, *Borrelia burgdorferi* sensu stricto, *Borrelia bavariensis*, *Borrelia spielmanii, Borrelia bissettiae*, and *Borrelia lusitaniae -* are present in Europe and considered pathogenic for human (Eisen, 2020; Margos et al., 2022; Rauter and Hartung, 2005). In metropolitan France, the annual incidence of Lyme borreliosis for 2024 was estimated at 53 cases per 100,000 inhabitants (95% Confidence Interval (95 CI): 43 - 63) (Réseau Sentinelle 2024). Other tick-borne bacterial pathogens of medical relevance in Europe include for instance: i) *Anaplasma phagocytophilum*: it causes human granulocytic anaplasmosis. Clinical cases are scarce but occur in Europe (Matei et al., 2019). In France cases were reported in eastern France in 2002 and 2012 (Edouard et al., 2012; Koebel et al., 2012; Remy et al., 2003); ii) *Neoehrlichia mikurensis*, the agent of neoehrlichiosis (Silaghi et al., 2016). In France four patients were found PCR positive for *N. mikurensis* between 2010 and 2018 (Boyer et al., 2021), iii) *Borrelia miyamotoi*, a relapsing fever group spirochete. It causes a flu-like illness in humans (Cleveland et al., 2023). Human cases have been identified in France (Franck et al., 2020a). This study has been the subject of a commentary and a response (Franck et al., 2020b; Wagemakers et al., 2020). All these pathogens are supposed to be mainly transmitted by *I. ricinus*.

Furthermore, several *Rickettsia* species also cause rickettsioses, such as *Rickettsia conorii*, responsible for Mediterranean spotted fever (MSF) and transmitted by *Rhipicephalus sanguineus* sensu lato ticks. Other species, including *Rickettsia helvetica*, *Rickettsia massiliae*, and *Rickettsia aeschlimannii*, transmitted by *I. ricinus*, *R. sanguineus* s.l. and *Hyalomma marginatum* respectively, can cause MSF-like syndromes, with fever, rash, and muscle aches being common symptoms, with more severe cases reported in Europe (Oteo and Portillo, 2012). Additionally, *Rickettsia slovaca* and *Rickettsia raoultii*, vectored by *Dermacentor marginatus* and *Dermacentor reticulatus*, are linked to SENLAT syndrome (Scalp Eschar and Neck LymphAdenopathy after Tick bite) which is characterized by scalp eschars and neck lymphadenopathy following tick bites (Oteo and Portillo, 2012).

Among protozoan pathogens, *Babesia* species, also transmitted by *I. ricinus* and *Dermacentor* species, cause babesiosis. While this disease primarily affects domestic and wild animals, human infections do occur. In Europe, *Babesia divergens*, *Babesia venatorum*, and *Babesia microti* are recognized as human pathogens (Hildebrandt et al., 2013). In France, since 1998, nine cases of babesiosis due to *B. divergens* have been reported, and two imported cases of *B. microti* (Hildebrandt et al., 2021).

Invasive tick species, such as *Hyalomma marginatum* and *Hyalomma lusitanicum* in France, can also be of public health importance. *H. marginatum* ticks indeed transmit Crimean-Congo hemorrhagic fever (CCHF) (Whitehouse, 2004), caused by the CCHF virus, which shows a high fatality rate in case of human infection. Although no human cases have been reported in France, the virus was recently isolated from *H. marginatum* ticks in the country (Bernard et al., 2024; Kiwan et al., 2024).

Environmental changes and human activities have probably influenced tick distribution and abundance in France. Ticks indeed occupy diverse habitats, increasing the risk of human exposure (Kahl and Gray, 2023). Monitoring the distribution and pathogen carriage of ticks is essential to assess public health risks and guide preventive actions. Traditional approaches, such as flagging or dragging, allow the collection of questing ticks and subsequent pathogen screening. However, these methods are labor-intensive and generally provide partial and piecewise information on the actual distribution of human exposure (Eisen and Eisen, 2016).

In contrast, citizen-science programs offer a practical and large-scale approach to assessing human-tick interactions (Eisen and Eisen, 2021). Such initiatives not only yield valuable data but also enhance public engagement awareness. In the United States, citizen science studies have successfully used human-biting ticks to monitor pathogen prevalence (Nieto et al., 2018; Porter et al., 2021, 2019). Similar initiatives were conducted in European countries such as Finland (Laaksonen et al., 2017) and Belgium (Lernout et al., 2019), with the latter focusing specifically on human-biting ticks as a direct measure of human exposure. In France, the CiTIQUE program has been established since 2017 to improve knowledge on tick ecology and tick-borne disease risks and to improve prevention strategies (Frey-Klett et al., 2018) This program allow citizens to report tick bites and send collected ticks to a national tick bank for research and surveillance purposes (Moutailler et al., 2023).

In this article, we aimed to characterize the tick species biting humans in France and to investigate the diversity and distribution of bacterial and protozoan microorganisms carried by these ticks. Some of these microorganisms can be pathogenic for humans. During the initial development of the program, we characterized a sub-sample of 2,009 human-biting ticks submitted to CiTIQUE to explore human exposure to tick species and tick-borne pathogens across France. The results are largely consistent with current knowledge, strengthening the relevance of such a program. They are discussed in the perspective of potential future improvement of the such citizen science programs.

## Materials and methods

### Tick surveillance, sample collection and identification

Ticks were submitted by citizen volunteers who chose to send tick specimens, in addition to reporting a tick bite to the CiTIQUE project via the “Signalement TIQUE” app, the website or a paper form (https://www.citique.fr/signalement-tique/). When reporting, citizens complete a questionnaire, which includes the following information: age, sex, location of the bite on the body, number of ticks attached, date of the bite (with an indication on the precision of the date), geographical location of the tick bite (with an indication on the precision of the GPS data), type of environment of the bite (e.g. forest, private yard…), type of activity that was performed at the time of the bite (e.g. sports, hiking…). Animal tick bite can also be reported through a different form. Citizens are informed they will not receive any individual results from the specimen’s analysis but that they can consult global results on the CiTIQUE website. Tick bite reporting and tick specimen collection began from the start of the program (July 2017) and is still ongoing.

Out of 17,010 human tick bite reports received from July 2017 to 2019, 4,776 were associated with one or more tick specimens. Tick sampling and analysis were initially carried out on a first-come, first-served basis, by selecting randomly only one tick per report (each report correspond to a different day and/or location). In 2017, only 191 reports with ticks were received, and 10 out of 13 administrative French regions (NUTS 1) having fewer than 20 reports (Table 1). The Nomenclature of territorial units for statistics (NUTS) was used in this study. It is a European classification used to reference countries’ regions for statistical purposes (https://ec.europa.eu/eurostat/web/nuts/). NUTS1 represents the larger level and the major socio-economic regions. We aimed at getting at least 150 ticks per region NUTS1 in order to get a spatial representation. As this number was only obtained for two regions (NAQ and GES) in 2018, some of the ticks received in 2019 were used to complete the sampling. It was not possible to obtain the required number of ticks for the PAC and 20R regions. A total of 2,009 ticks was finally sampled between 2017 to 2019 (Table 1).

**Table 1.** Sample distribution of human-biting ticks by year and region with cumulative number over the years. Regions: 20R: Corse; ARA: Auvergne-Rhône-Alpes; BFC: Bourgogne-Franche-Comté; BRE: Bretagne; CVL: Centre-Val de Loire; GES: Grand-Est; HDF; Hauts-De-France; IDF: Île-de-France; NOR: Normandie; NAQ: Nouvelle-Aquitaine; OCC: Occitanie; PAC: Provence-Alpes-Côte d’Azur; PDL: Pays-de-la-Loire.

| Region | In 2017 | In 2017+2018 | In 2017+2018+2019 |
| --- | --- | --- | --- |
| **France** | **191** | **1,422** | **2,009** |
| **GES** | 49 | 208 | 298 |
| **BRE** | 12 | 122 | 224 |
| **NAQ** | 15 | 153 | 174 |
| **ARA** | 31 | 138 | 166 |
| **PDL** | 6 | 71 | 166 |
| **HDF** | 5 | 96 | 159 |
| **BFC** | 14 | 145 | 157 |
| **IDF** | 19 | 142 | 156 |
| **NOR** | 7 | 74 | 150 |
| **CVL** | 5 | 114 | 149 |
| **OCC** | 28 | 126 | 148 |
| **PAC** | 0 | 31 | 59 |
| **20R** | 0 | 2 | 3 |

The CiTIQUE program grew in term of people and associated skills during this study, resulting in different treatments of ticks regarding their identification. At first, no one was competent in tick morphological identification and we relied only on molecular identification with the high-throughput microfluidic real-time PCR method which could only identify three species including *Ixodes ricinus*, *Dermacentor marginatus* and *Dermacentor reticulatus* (Fig. 1.), without developmental stage identification. Tick species, targeted genes and primers sets are listed in Table S1 (supplementary material).

**Fig. 1.**
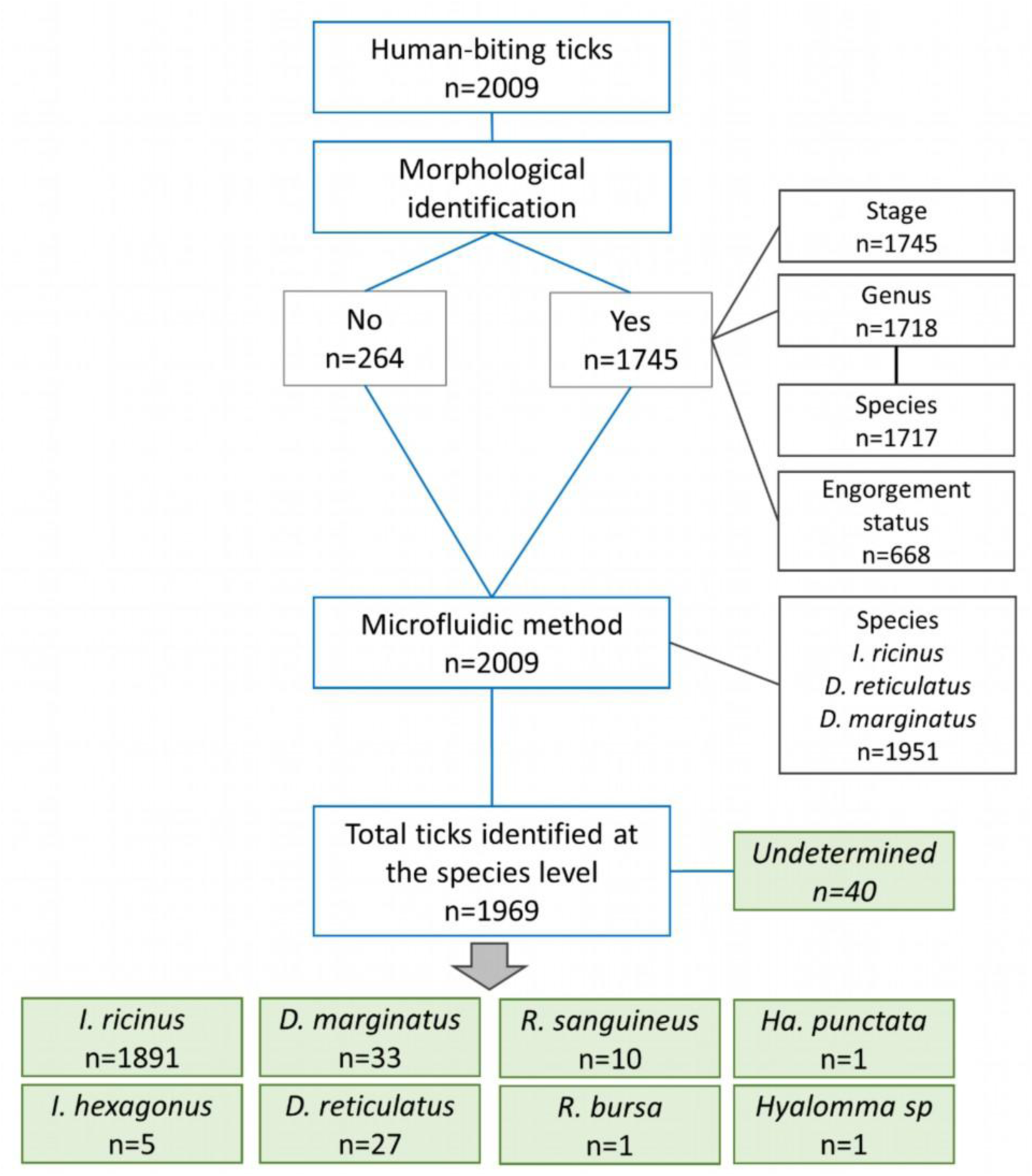
Flow diagram of performed analyses on human-biting ticks in this study. *I., Ixodes; D., Dermacentor; R., Rhipicephalus; Ha., Haemaphysalis*

After implementing morphological tick identification, we started to collect additional data on tick engorgement, as a rough indicator of tick feeding time. Ticks were determined as engorged when body modification characteristics of blood engorgement were observed, which appear after more than 24h of feeding (Yeh et al., 1995). This characterization was made to separate ticks that were removed rapidly (less than 24h) from ticks that were retrieved at a later time, increasing the risk of transmission of a pathogen if the tick was infected. Out of 264 ticks that were not identified for developmental stage, 13 were not identified by molecular method either, because they belong to species other than *Ixodes ricinus* or *Dermacentor reticulatus* or *Dermacentor marginatus*; 27 ticks were also identified for developmental stage but not for the species by molecular method. Morphological identification of ticks was performed using two identification keys (Estrada-Peña et al., 2018; Pérez-Eid, 2007). Out of 1745 ticks identified morphologically, 668 had their engorgement status recorded (Fig. 1.).

### Detection of microorganisms

#### DNA extraction

Ticks were homogenised using 2.8 mm stainless steel beads in a Precellys 24 lyser/homogeniser (Bertin, France) at 5500 rpm for 20s. DNA from single specimens was extracted using NucleoSpin® Tissue kit (Macherey-Nagel, Germany), following manufacturer’s instructions with an exception: elution was performed in 50µl with BE buffer heated at 70°C.

#### DNA pre-amplification and high-throughput microfluidic real-time PCR

Microorganisms, targeted genes and primers sets are listed in Table S1 (supplementary material). Every species of a genus or a family of microorganisms are not researched. Consequently, it is possible to detect only the presence of a microorganism at the genus level (i.e. *Rickettsia* spp.) without having a corresponding species detected. Some of the targeted microorganisms are known to be pathogenic for humans, some are not. These differences, along the associated potential vector tick species, are summarized in Table S2 (supplementary material).

A pre-amplification step by PCR with the Preamp Master Mix (Standard BioTools, USA) was performed on the samples according to Melis et al (2024) in order to enhance the sensitivity of microfluidic PCR for microorganism detection. All the primers used for microorganism detection (Table S1) were pooled, at a final concentration of 0.2 μM each. The reaction was carried out as described in Melis et al (2024). Pre-amplified samples were diluted 1:10 and then stored at - 20 °C until further analysis.

High-throughput microfluidic real-time PCR was performed using a BioMark™ real-time PCR system (Standard BioTools, USA), and the 48.48 Dynamic Array™ (Standard BioTools, USA). The chip includes two sets of 48 inlets, allowing the simultaneous loading of 48 samples and 48 assays reaction mix (with specific primers and probe). All 2,304 (48*48) sample-assay combinations were processed simultaneously by the automated microfluidic system (see Michelet et al., 2014; Melis et al., 2024 for details). Amplifications were performed as in Melis et al (2024).

#### Validation of the results by PCR and DNA sequencing

In order to evaluate, refine and validate the results obtained with the BioMark™ system, conventional PCRs were performed on DNA extracts from a subset of representative positive samples following Melis et al (2024). This representative subset included ticks in which *Borrelia* spp.*, Rickettsia* spp. or *A. phagocytophilum* were detected. This representative subset included at least 20 % of all positives to each microorganism. For validation of the microfluidic real-time PCR assays, only samples showing low Cq values (<30) were subjected to conventional PCR assays. Different primers were used for each microorganism compared to those in the microfluidic real-time PCR, following published thermal protocols Melis et al (2024). The amplicons were bidirectionally sequenced by Eurofins (Germany), and then sequencing results were inspected and processed using the BioEdit software (Ibis Biosciences, Carlsbad). The results were compared with the NCBI nucleotide database using BLASTN.

#### Statistical and spatial analysis

The dataset presents information on the 2,009 ticks analyzed for microorganism detection and identification. The 72 fields are grouped into five categories: description of the tick bite declarations (eight fields), tick characterization (12 fields), microorganism detection with microfluidic method (44 fields), microorganism detection by sequencing (two fields), synthesis from microfluidic assay results (six field). Dataset is available at the Recherche Data Gouv Repository:

Infection rates were calculated as the proportion of infected ticks among the total number of ticks per species or region. The corresponding 95% confidence intervals (95 CI) were estimated using a binomial test (binom.test function in R).

The association between infection status and tick species, developmental stage, and engorgement status was assessed using a binomial logistic regression (logit link). The same modelling approach was applied to test whether coinfection status varied with species or developmental stage.

To assess whether infection rates differed among regions, we first conducted an exploratory clustering based on observed regional infection rates. We applied hierarchical agglomerative clustering using complete linkage on Euclidean distances, which allowed us to identify groups of regions with similar infection rate levels.

We then statistically evaluated differences among these clusters by fitting a binomial logistic regression, modelling the number of infected ticks over the total sampled in each region and using the cluster assignments as a categorical predictor. These analyses were conducted only for *Borrelia burgdorferi* sensu lato and *Rickettsia helvetica*, as these were the only taxa with sufficient observations across French regions.

When models had significant predictors, odds ratios were derived from post-hoc pairwise comparisons using estimated marginal means (emmeans package v2.0.0; Lenth et al., 2025) with Tukey adjustment to control the family-wise error rate.

All statistical analyses and spatial mapping were performed using R version 4.5.1 (R Core Team, 2025).

## Results

### Tick stages and species distribution

Of 2,009 ticks submitted by citizens, 1,745 were morphologically identified to determine their developmental stage (larva, nymph, adult female or male), and 1,717 were identified taxonomically to the genus and species level. All ticks were tested with the microfluidic chip assay, allowing to confirm morphological taxonomic identification for the following species: *I. ricinus*, *D. marginatus* and *D. reticulatus*. Only four ticks were not morphologically confirmed as *I. ricinus*. Among the 1,745 ticks identified for developmental stage, nymphs were the most represented (1346; 77.1%; 95 CI: 75.1 - 79.1 - the numbers correspond to the number of individuals, the percentage of individuals and its 95% confidence interval, respectively), followed by adult females (272; 15.6%; 95 CI: 13.9 - 17.4), larvae (112; 6.4%; 95 CI: 5.3 - 7.7), and adult males (15; 0.9%; 95 CI: 0.5 – 1.4). Among the 2,009 ticks, five genera and seven species were identified (Fig. Fig. 1, Fig. 2, Table S3). The most frequent species was *I. ricinus* (1,891 ticks). We also identified five *I. hexagonus* leading to 94.4% *Ixodes* ticks in the dataset. In addition, 60 *Dermacentor* ticks were identified (33 *D. marginatus* and 27 *D. reticulatus)* with 10 nymphs (nine *D. marginatus* and one *D. reticulatus*) and 50 adults, along with 11 *Rhipicephalus* ticks (10 *R. sanguineus* s.l. and one *R. bursa)*, one *Haemaphysalis punctata* and one *Hyalomma* spp tick.

**Fig. 2.**
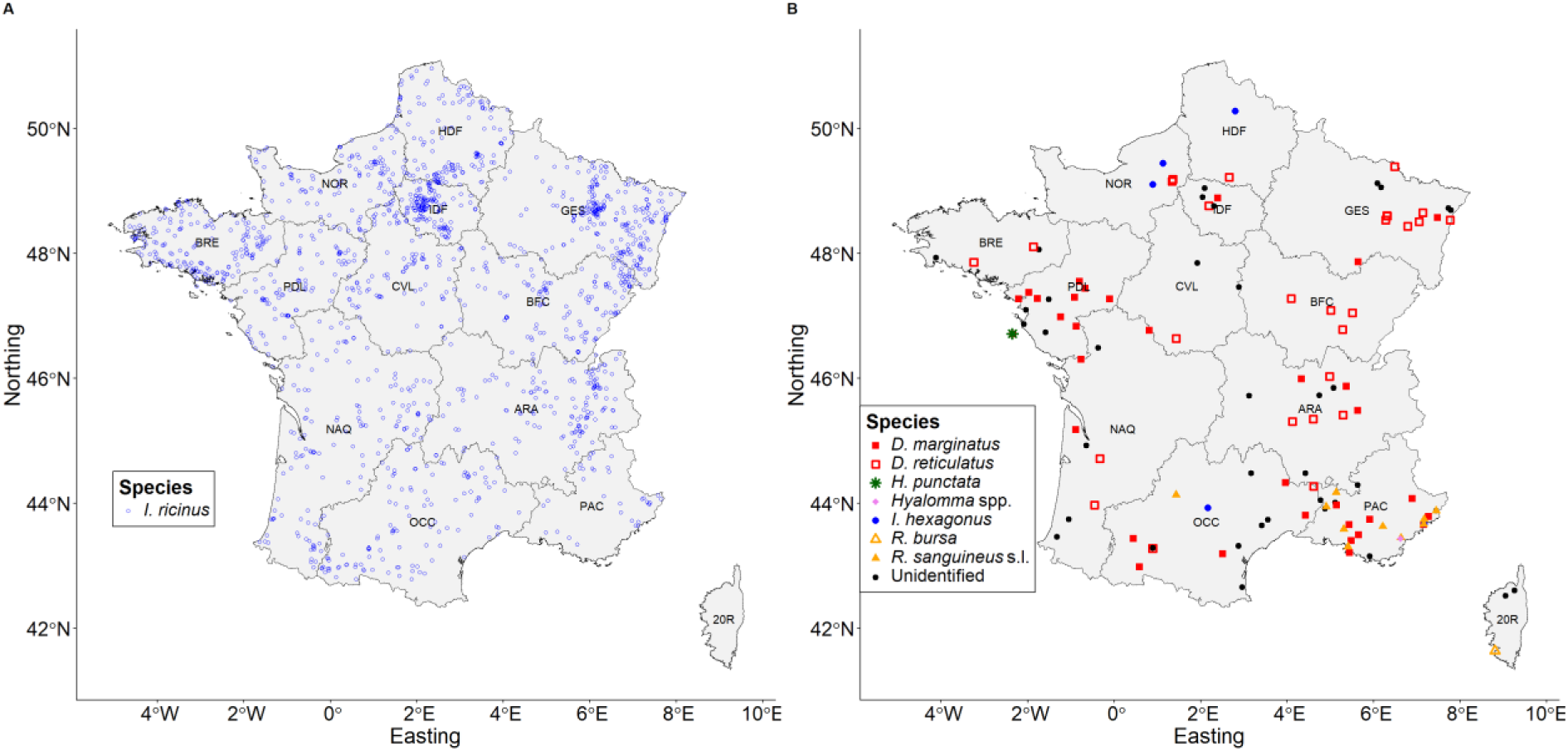
Geographic distribution of the 2,009 human-biting ticks studied, differentiated by tick species (A. *Ixodes ricinus* distribution; B. Other tick species distribution). Region: 20R: Corse; ARA: Auvergne-Rhône-Alpes; BFC: Bourgogne-Franche-Comté; BRE: Bretagne; CVL: Centre-Val de Loire; GES: Grand-Est; HDF; Hauts-De-France; IDF: Île-de-France; NOR: Normandie; NAQ: Nouvelle-Aquitaine; OCC: Occitanie; PAC: Provence-Alpes-Côte d’Azur; PDL: Pays-de-la-Loire. Sources: base map GADM.

Fig. shows the distribution of the different tick species collected and selected across France. *I. ricinus*, the predominant tick species, was sampled in almost all regions. It accounted for 88% to 99% of ticks in most areas. Notable exceptions were Corse region (20R), where no *I. ricinus* were found among the three collected ticks, and the Provence-Alpes-Côte d’Azur region (PAC), where it represented only 61% of identified ticks. In the PAC region, *D. marginatus* and *R. sanguineus* s.l. each accounted for 15% of the identified ticks, reflecting a more diverse tick community in this area. *Dermacentor marginatus* was detected in seven out of twelve regions (ARA, GES, IDF, NAQ, OCC, PAC, and PDL), but was absent from samples of the northern and western regions of the country, including BRE, NOR, HDF, CVL, and BFC. On the other hand, *D. reticulatus* was identified in all regions except for PDL, PAC and 20R. Ticks of the genus *Rhipicephalus* were identified in the southern regions of France, with nine *R. sanguineus* s.l. ticks collected in PAC, one in OCC, and one *R. bursa* tick in 20R. The identified *Hyalomma* tick was collected in the PAC region. The *Haemaphysalis punctata* tick was collected in the Pays de la Loire region (PDL), on an Atlantic Island. Five *I. hexagonus* ticks were identified across four regions: two in the north, Hauts-de-France and Normandy (HDF and NOR), and two in the south, Provence-Alpes-Côte d’Azur and Occitanie (PAC and OCC).

Of the 668 ticks for which engorgement status was recorded, 292 (44%; 95 CI: 40 - 48) were reported as engorged. Engorgement status varied by developmental stage and sex: 249 of 519 nymphs (48%; 95 CI: 44 - 52), 30 of 109 females (28%; 95 CI: 19 - 37) were engorged, only one male of 13, and 12 of 27 larvae (44%; 95 CI: 25 - 65). Engorgement status varied significantly across developmental stages (Deviance = 24.401, p-value <0.001). Post-hoc pairwise comparisons showed that nymphs were significantly more often engorged than females with odds of being fed 2.42 higher (OR = 2.4; 95 CI: 1.25 - 4.48, p-value = 0.0029).

Regarding species, no significant association was found with engorgement status (Deviance = 0.68, p-value = 0.41). Engorgement was observed in 278 of 624 (45%; 95 CI: 41- 49) *I. ricinus*, four of five (80%; 95 CI: 28 - 99) *I. hexagonus*, six of 15 (40%; 95 CI: 16 - 68) *D. marginatus*, and four of 11 (36%; 95 CI: 11 - 69) *D. reticulatus* individuals. No engorged ticks were recorded for *Ha. punctata*, *Hyalomma* spp., *R. bursa*, or *R. sanguineus*.

### Validation of the results obtained with the BioMark™ system

For *Borrelia* spp., on the positive ticks with chip method, 105 (20%) were tested for confirmation using flaB gene amplification (Melis et al., 2024; Michelet et al., 2014), and 61 sequences were obtained for different species. In total, 309 samples were then validated for *Borrelia* detection after extrapolation.

For *Rickettsia* spp., on the positive ticks with chip method, 71 (20%) were tested for confirmation using gltA or ompB genes amplification (Melis et al., 2024), and 35 sequences were obtained. In total, 172 samples were validated for *Rickettsia* detection after extrapolation.

For *A. phagocytophilum*, among 142 positive ticks with the chip method, 106 were tested for confirmation. The samples were analyzed by qPCR using the taqman system targeting a 122 bp fragment of the *A. phagocytophilum* msp2 gene (Drazenovich et al., 2006) with Biorad’s SoAdvanced™ Universal Probes Supermix qPCR mix. The 14 unconfirmed samples were also at the detection limit of the microchip method (high CT). In total, the 142 samples were validated for *A. phagocytophilum* infection after extrapolation.

### Microorganisms identification

The high-throughput PCR method was applied to each of the 2,009 ticks, leading to the detection of 26 different microorganisms in 618 ticks (infection rate: 30.8%; 95 CI: 28.7 - 32.8; Table 2, Table S4 and S5), including 18 human pathogens in 556 ticks (infection rate :27.7%; 95 CI: 25.7 – 29.7. Among the pathogen-infected ticks, 90 ticks (4.5%; 95 CI: 3.6 - 5.5; Table 2) were infected by at least two different pathogens.

**Table 2.**
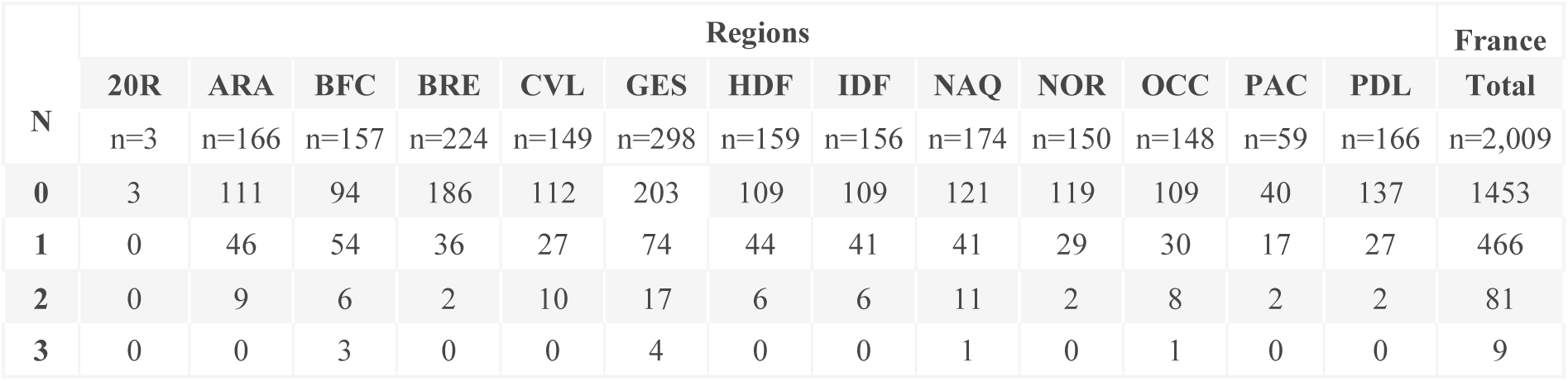
Number of human-biting ticks carrying N human pathogens per region. N=Number of pathogens found in ticks. n: total number of ticks in each region. Region: 20R: Corse; ARA: Auvergne-Rhône-Alpes; BFC: Bourgogne-Franche-Comté; BRE: Bretagne; CVL: Centre-Val de Loire; GES: Grand-Est; HDF: Hauts-de-France; IDF: Île-de-France; NAQ: Nouvelle-Aquitaine; NOR: Normandie; OCC: Occitanie; PAC: Provence-Alpes-Côte d’Azur; PDL: Pays-de-la-Loire.

**Table 1.** Number of human-biting ticks carrying pathogenic (to humans) and non-pathogenic microorganisms, with infection rates, per tick species. n: total number of ticks analyzed for each species. Results: infected tick number; infection rate in percentage (95% confidence interval). Genera of microorganisms in bold letters. Non-pathogenic microorganisms are marked by an asterisk *.

| Microorganisms | Tick species |  |  |  |  |
| --- | --- | --- | --- | --- | --- |
|  | *D. marginatus*<br>n=33 | *D. reticulatus*<br>n=27 | *I. ricinus*<br>n=1,891 | *R. sanguineus* s.l.<br>n=10 | Unidentified<br>n=40 |
| ***Borrelia* spp.** | 0; 0%<br>(0-10.6) | 0; 0%<br>(0-12.8) | 309; 16.3%<br>(14.7-18.1) | 0; 0%<br>(0-30.8) | 0; 0%<br>(0-8.6) |
| ***Borrelia burgdorferi* s.l.** | 0; 0%<br>(0-10.6) | 0; 0%<br>(0-12.8) | 291; 15.4%<br>(13.8-17.1) | 0; 0%<br>(0-30.8) | 0; 0%<br>(0-8.6) |
| *Borrelia afzelii* | 0; 0%<br>(0-10.6) | 0; 0%<br>(0-12.8) | 136; 7.2%<br>(6.1-8.5) | 0; 0%<br>(0-30.8) | 0; 0%<br>(0-8.6) |
| *Borrelia garinii* | 0; 0%<br>(0-10.6) | 0; 0%<br>(0-12.8) | 80; 4.2%<br>(3.4-5.2) | 0; 0%<br>(0-30.8) | 0; 0%<br>(0-8.6) |
| *Borrelia valaisiana* * | 0; 0%<br>(0-10.6) | 0; 0%<br>(0-12.8) | 37; 2%<br>(1.4-2.7) | 0; 0%<br>(0-30.8) | 0; 0%<br>(0-8.6) |
| *Borrelia burgdorferi* s.s. | 0; 0%<br>(0-10.6) | 0; 0%<br>(0-12.8) | 25; 1.3%<br>(0.9-1.9) | 0; 0%<br>(0-30.8) | 0; 0%<br>(0-8.6) |
| *Borrelia spielmanii* | 0; 0%<br>(0-10.6) | 0; 0%<br>(0-12.8) | 8; 0.4%<br>(0.2-0.8) | 0; 0%<br>(0-30.8) | 0; 0%<br>(0-8.6) |
| *Borrelia lusitaniae* | 0; 0%<br>(0-10.6) | 0; 0%<br>(0-12.8) | 5; 0.3%<br>(0.1-0.6) | 0; 0%<br>(0-30.8) | 0; 0%<br>(0-8.6) |
| *Borrelia miyamotoi* | 0; 0%<br>(0-10.6) | 0; 0%<br>(0-12.8) | 25; 1.3%<br>(0.9-1.9) | 0; 0%<br>(0-30.8) | 0; 0%<br>(0-8.6) |
| ***Anaplasma* spp.** | 2; 6.1% | 1; 3.7% | 135; 7.1% | 1; 10% | 3; 7.3% |
|  | **(0.7-20.2)** | **(0.1-19)** | **(6-8.4)** | **(0.3-44.5)** | **(1.5-19.9)** |
| *Anaplasma phagocytophilum* | 2; 6.1%<br>(0.7-20.2) | 1; 3.7%<br>(0.1-19) | 135; 7.1%<br>(6-8.4) | 1; 10%<br>(0.3-44.5) | 3; 7.3%<br>(1.5-19.9) |
| ***Ehrlichia* spp.** | **0; 0%**<br>**(0-10.6)** | **0; 0%**<br>**(0-12.8)** | **54; 2.9%**<br>**(2.2-3.7)** | **0; 0%**<br>**(0-30.8)** | **3; 7.3%**<br>**(1.5-19.9)** |
| *Neoehrlichia mikurensis* | 0; 0%<br>(0-10.6) | 0; 0%<br>(0-12.8) | 49; 2.6%<br>(1.9-3.4) | 0; 0%<br>(0-30.8) | 1; 2.4%<br>(0.1-12.9) |
| ***Rickettsia* spp.** | **14; 42.4%**<br>**(25.5-60.8)** | **13; 48.1%**<br>**(28.7-68.1)** | **134; 7.1%**<br>**(6-8.3)** | **3; 30%**<br>**(6.7-65.2)** | **10; 24.4%**<br>**(12.4-40.3)** |
| *Rickettsia helvetica* | 0; 0%<br>(0-10.6) | 0; 0%<br>(0-12.8) | 120; 6.3%<br>(5.3-7.5) | 0; 0%<br>(0-30.8) | 0; 0%<br>(0-8.6) |
| *Rickettsia raoultii* | 4; 12.1%<br>(3.4-28.2) | 13; 48.1%<br>(28.7-68.1) | 8; 0.4%<br>(0.2-0.8) | 0; 0%<br>(0-30.8) | 0; 0%<br>(0-8.6) |
| *Rickettsia slovaca* | 6; 18.2%<br>(7-35.5) | 0; 0%<br>(0-12.8) | 1; 0.1%<br>(0-0.3) | 0; 0%<br>(0-30.8) | 1; 2.4%<br>(0.1-12.9) |
| *Rickettsia massiliae* | 0; 0%<br>(0-10.6) | 0; 0%<br>(0-12.8) | 0; 0%<br>(0-0.2) | 2; 20%<br>(2.5-55.6) | 1; 2.4%<br>(0.1-12.9) |
| *Rickettsia monacensis* | 0; 0%<br>(0-10.6) | 0; 0%<br>(0-12.8) | 1; 0.1%<br>(0-0.3) | 0; 0%<br>(0-30.8) | 0; 0%<br>(0-8.6) |
| *Rickettsia aeschlimannii* | 0; 0%<br>(0-10.6) | 0; 0%<br>(0-12.8) | 0; 0%<br>(0-0.2) | 0; 0%<br>(0-30.8) | 2; 4.9%<br>(0.6-16.5) |
| *Rickettsia felis* * | 0; 0%<br>(0-10.6) | 0; 0%<br>(0-12.8) | 0; 0%<br>(0-0.2) | 0; 0%<br>(0-30.8) | 1; 2.4%<br>(0.1-12.9) |
| ***Bartonella* spp.** | **0; 0%**<br>**(0-10.6)** | **0; 0%**<br>**(0-12.8)** | **1; 0.1%**<br>**(0-0.3)** | **0; 0%**<br>**(0-30.8)** | **0; 0%**<br>**(0-8.6)** |
| *Bartonella henselae* | 0; 0%<br>(0-10.6) | 0; 0%<br>(0-12.8) | 1; 0.1%<br>(0-0.3) | 0; 0%<br>(0-30.8) | 0; 0%<br>(0-8.6) |
| *Francisella-like endosymbiont* * <sup>&</sup> | 1; 3%<br>(0.1-15.8) | 26; 96.3%<br>(81-99.9) | 23; 1.2%<br>(0.8-1.8) | 0; 0%<br>(0-30.8) | 3; 7.3%<br>(1.5-19.9) |
| ***Apicomplexa*** | **1; 3%**<br>**(0.1-15.8)** | **1; 3.7%**<br>**(0.1-19)** | **28; 1.5%**<br>**(1-2.1)** | **0; 0%**<br>**(0-30.8)** | **3; 7.3%**<br>**(1.5-19.9)** |
| ***Babesia* spp.** | **0; 0%**<br>**(0-10.6)** | **1; 3.7%**<br>**(0.1-19)** | **24; 1.3%**<br>**(0.8-1.9)** | **0; 0%**<br>**(0-30.8)** | **0; 0%**<br>**(0-8.6)** |
| *Babesia venatorum* | 0; 0%<br>(0-10.6) | 0; 0%<br>(0-12.8) | 14; 0.7%<br>(0.4-1.2) | 0; 0%<br>(0-30.8) | 0; 0%<br>(0-8.6) |
| *Babesia microti* | 0; 0%<br>(0-10.6) | 0; 0%<br>(0-12.8) | 5; 0.3%<br>(0.1-0.6) | 0; 0%<br>(0-30.8) | 0; 0%<br>(0-8.6) |
| *Babesia divergens* | 0; 0%<br>(0-10.6) | 1; 3.7%<br>(0.1-19) | 4; 0.2%<br>(0.1-0.5) | 0; 0%<br>(0-30.8) | 0; 0%<br>(0-8.6) |
| *Babesia canis* * | 0; 0%<br>(0-10.6) | 0; 0%<br>(0-12.8) | 1; 0.1%<br>(0-0.3) | 0; 0%<br>(0-30.8) | 0; 0%<br>(0-8.6) |
| ***Theileria* spp.\*** | 0; 0%<br>(0-10.6) | 0; 0%<br>(0-12.8) | 0; 0%<br>(0-0.2) | 0; 0%<br>(0-30.8) | 2; 4.9%<br>(0.6-16.5) |
| ***Hepatozoon* spp.\*** | **1; 3%**<br>**(0.1-15.8)** | **0; 0%**<br>**(0-12.8)** | **2; 0.1%**<br>**(0-0.4)** | **0; 0%**<br>**(0-30.8)** | **0; 0%**<br>**(0-8.6)** |

1 <sup>&</sup>Total of *Francisella*-like endosymbiont type 1 and 2

Sequences for *Borrelia* and *Rickettsia* species were deposited on Genbank (*B. afzelii*: PX692869; *B*. *burgdorferi* ss: PX692870, PX692871, PX692872; *B*. *garinii*: PX692873, PX692874, PX692875, PX692876; *B*. *valaisiana*: PX692877, PX692878; *B*. *lusitaniae*: PX692879, PX692880; *B*. *spielmanii*: PX692881; *R*. *raoultii*: PX692882, PX692883; *R*. *slovaca*: PX692884, PX692885; *R*. *monacensis*: PX692886; *R*. *aeschlimannii*: PX692887; *R*. *massiliae*: PX692888, PX692889).

No significative difference was observed in infection with at least one microorganism for both engorgement status or species (respectively, Deviance = 0.0679, p-value = 0.79; Deviance = 10.4516, p-value = 0.16).

Development stage had a significant effect (Deviance = 16.1447, p-value = 0.001). However, post-hoc pair-wise comparisons revealed no significant pairwise differences between stages except between larvae and males: infection probability was 15 times higher in males relative to larvae (OR = 15, 95 CI: 1.18 - 203, p-value = 0.031). Given the very small number of male ticks (n=15), this result carries substantial uncertainty. Among the 1,891 *I. ricinus* ticks, 291 (15.4%; 95 CI: 14.7 - 18.1; Table 3) were infected with *B. burgdorferi* s.l. and 25 (1.3%; 95 CI: 0.9 – 1.9; Table 3) with *Borrelia miyamotoi*. In addition, *A. phagocytophilum* was detected in 135 ticks (7.1%; 95 CI: 6.0 - 8.4; Table 3), while 134 (7.1%; 95 CI: 6.0 - 8.3; Table 3) were positive for *Rickettsia* spp., 54 (2.9%; 95 CI: 2.2 – 3.7; Table 3) for *Ehrlichia* spp. and 24 (1.3%; 95 CI:0.8 – 1.9; Table 3) for *Babesia* spp. None of the five *I. hexagonus* ticks tested positive for any of the screened pathogens.

Among the 60 *Dermacentor* ticks, *A. phagocytophilum* was detected in one *D. marginatus* and two *D. reticulatus* ticks. Fourteen *D. marginatus* ticks (42%; 95 CI: 25.5 – 60.8; Table 3) and 13 *D. reticulatus* ticks (48%; 95 CI: 28.7 – 68.1; Table 3) were infected with *Rickettsia* spp. Four out of 10 *R. sanguineus* s.l. ticks were infected: one with *A. phagocytophilum* and three with *Rickettsia* spp. The single *Hyalomma* spp. tick was positive for *Rickettsia* spp. No pathogens were detected in the identified *R. bursa* tick or in the *H. punctata* tick (Table 3).

### Borrelia species

*Borrelia* spp. was detected in 309 *I. ricinus* ticks, corresponding to an infection rate of 16.3% (95 CI: 14.7-18.1; Table 3). These detections fall into two different species complexes, one that includes the agent of human Lyme borreliosis (*B. burgdorferi* s.l.) and the other, the agents of the human relapsing fever (more specifically *B. miyamotoi* here). Infection rates varying by region and *Borrelia* species.

Across all developmental stages, the average *B. burgdorferi* s.l. infection rate in *I. ricinus* ticks in France was 15.4% (291/1891; 95 CI: 13.8 -17.1; Table 3, Fig. 3), ranging from 2.9% (1/35; 95 CI: 0.07 – 14.9) in the PAC region to 21.1% (60/284; 95 CI: 16.5 – 26.3) in the GES region.

**Fig. 3.**
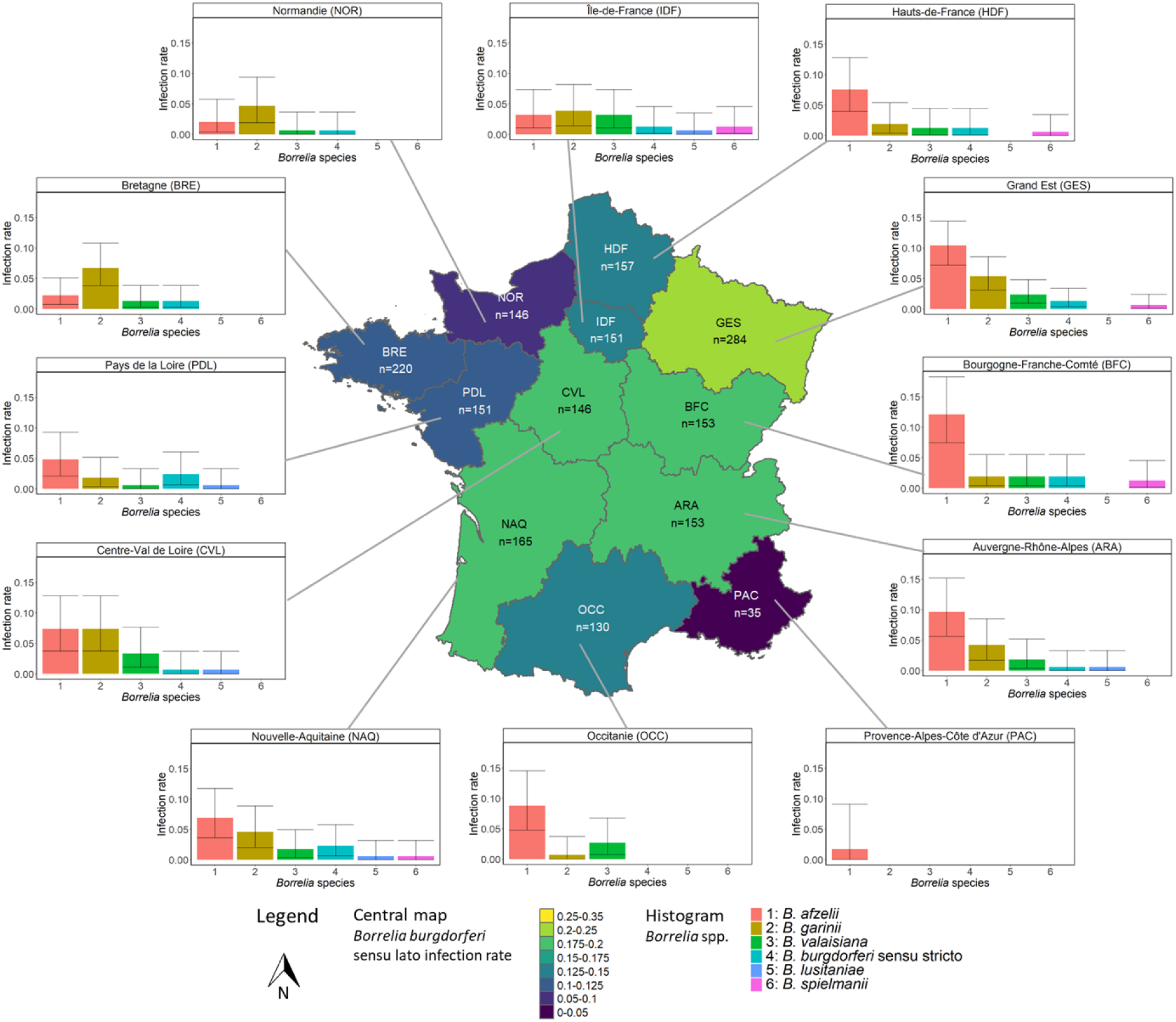
Infection rates of *Borrelia burgdorferi* sensu lato per region in analyzed human-biting *Ixodes ricinus* ticks (central map), and histograms of the specific infection rate of the main *Borrelia* species in each region. Central map: n: number of *I. ricinus* ticks analyzed in the considered region. Histogram: Confidence interval 95%. Region: 20R: Corse; ARA: Auvergne-Rhône-Alpes; BFC: Bourgogne-Franche-Comté; BRE: Bretagne; CVL: Centre-Val de Loire; GES: Grand-Est; HDF: Hauts-de-France; IDF: Île-de-France; NAQ: Nouvelle-Aquitaine; NOR: Normandie; OCC: Occitanie; PAC: Provence-Alpes-Côte d’Azur; PDL: Pays-de-la-Loire.

The hierarchical clustering on *B. burgdorferi* s.l. infection rate identified four regional groups. Cluster 1, a high-infection rate group (infection rate >15.4%), included ARA, BFC, CVL, GES and NAQ. Cluster 2, with moderate infection rate (10–15%), comprised BRE, HDF, IDF and OCC. Cluster 3, a low-to-moderate group (8–10.24%), included PDL and NOR. Cluster 4 showed near-zero infection rates and included 20R and PAC. Cluster 1 exhibited significantly higher infection rates than Clusters 2, 3 and 4 (OR = 1.62, p-value = 0.004; OR = 2.27, p-value < 0.001; and OR = 13.98, p-value = 0.045, respectively). No significant differences were detected among the remaining clusters.

Infection rate was 18.8% in adult females (41/218, 95 CI: 13.8 – 24.6), followed by nymphs 15.1% (200/1324, 95 CI: 13.2 – 17.1), larvae 7.3% (8/110, 95 CI: 3.2 – 13.8). One out of three adult male ticks was positive.

The following species of the *B. burgdorferi* s.l. complex (Fig. 3) were detected, listed in descending order of occurrence, with the corresponding number of *I. ricinus* ticks infected, the infection rate and 95% confidence interval: *B. afzelii* (136; 7.2%; 95 CI: 6.1 – 8.5), *B. garinii* (80; 4.2%; 95 CI: 3.4 – 5.3), *B. valaisiana* (37; 2.0%; 95 CI: 1.4 – 2.7), *B. burgdorferi* s.s. (25; 1.3%; 95 CI: 0.9 – 1.9), *B. spielmanii* (8; 0.4%; 95 CI 0.18 – 0.83), *B. lusitaniae* (5; 0.3%; 95 CI: 0.09 – 0.6).

### Rickettsia species

Out of 2,009 human-biting ticks tested, 172 were tested positive for *Rickettsia* spp., with 120 *R. helvetica,* the main species detected, all found in *I. ricinus* ticks. Other detected species included: *R. raoultii* (25 ticks, comprising 13 *D. reticulatus*, eight *I. ricinus* and four *D. marginatus*); *R. slovaca* (eight ticks: six *D. marginatus*, one *I. ricinus* and one undetermined tick); *R. massiliae* (three ticks: two *R. sanguineus* s.l. and one undetermined tick); *R. aeschlimannii* (two undetermined ticks); *R. monacensis* (one *I. ricinus* tick) and *R. felis* (one undetermined tick). *R. conorii* was not detected (Table 3).

As the high-throughput results for 171 *Rickettsia* were confirmed at the species level by PCR and sequencing, results allow the analysis of the variation of the number of species observed from one region to another (Fig. 4). Five *Rickettsia* species were detected in the south-east in PAC region (i.e. *R. helvetica, R. raoultii, R. aeschlimannii,* and *R. massiliae*) compared with just one in the CVL region (*R. helvetica*) in the north-west.

**Fig. 4.**
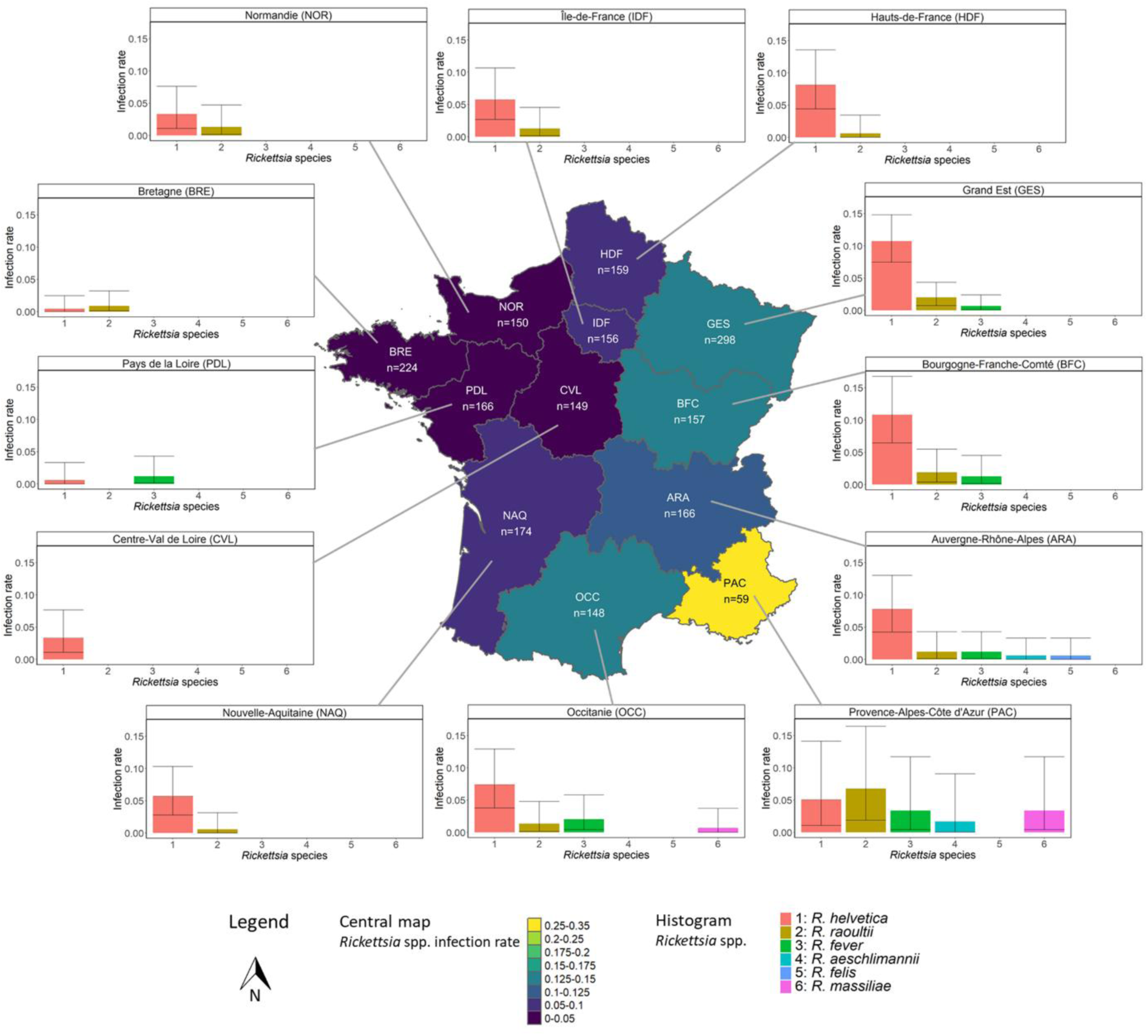
Infection rates of *Rickettsia* spp. per region in analyzed human-biting ticks (central map), and histogram of the specific infection rate of the main *Rickettsia* species in each region. Central map: n: number of ticks analyzed in the considered region. Histogram: Confidence interval 95%. Region: 20R: Corse; ARA: Auvergne-Rhône-Alpes; BFC: Bourgogne-Franche-Comté; BRE: Bretagne; CVL: Centre-Val de Loire; GES: Grand-Est; HDF: Hauts-de-France; IDF: Île-de-France; NAQ: Nouvelle-Aquitaine; NOR: Normandie; OCC: Occitanie; PAC: Provence-Alpes-Côte d’Azur; PDL: Pays-de-la-Loire.

*R. helvetica* was exclusively detected in *I. ricinus* ticks, with an average infection rate at the country scale of 6.3% (120/1891; 95 CI: 5.3 - 7.5; Table 3, Fig. 4).

The hierarchical clustering of *R. helvetica* infection rates identified three regional groups. Cluster 1, a high-infection rate group (>9%), comprised the eastern regions BFC and GES. Cluster 2, with moderate infection rate (6–9%), included centrally located regions (IDF, NAQ, HDF, PAC, ARA and OCC). Cluster 3, a low-infection rate group (<6%), comprised the western regions CVL, NOR, BRE and PDL. Cluster 3 showed significantly lower infection rates than both Cluster 1 and Cluster 2 (OR = 6.85, p-value < 0.001 and OR = 4.37, p-value < 0.001, respectively). Cluster 1 had higher infection rate than Cluster 2 but the relationship was not significant (OR = 1.57, p-value= 0.070).

*R. raoultii* was present in 25 ticks distributed throughout France, with the exception of three regions 20R, CVL and PDL. Infection rates were generally below 2%, except in the PAC region, where they reached 6.8% (4/59; 95 CI: 1.8 – 16.5) (Fig. 4).

### Anaplasma species

Of the six *Anaplasma* species targeted by the microfluidic chip, only *A. phagocytophilum* was detected, with 142 positive ticks. This corresponds to an average infection rate among French samples of 7.1% (142/2009; 95 CI: 6.0 - 8.4). Among the 142 positive ticks, the majority were *I. ricinus* (135/142, 95.1%; 95 CI: 90.1 – 98.0). The remaining seven positive ticks included three *Dermacentor* specimens (two *D. marginatus* in the ARA and PAC regions and one *D. reticulatus* in the NAQ region), one *R. sanguineus* s.l. from PAC region, and three undetermined ticks collected in IDF, NAQ and PDL regions. Infection rates in *I. ricinus* varied across regions, from 3.9% (11/284; 95 CI: 1.95 – 6.8) in GES region to 12.6% (19/151; 95 CI: 7.7 – 19.0) in IDF region (Fig. 5). Considering all genera and species of ticks, the infection rate was the highest for adult female ticks (25/272, 9.2%; 95 CI: 6.0 – 13.3) followed by nymphs (105/1346, 7.8%; 95 CI: 6.4 – 9.4) and larvae (5/112, 4.5%; 95 CI: 1.5 – 10.1). No males were infected.

**Fig. 5.**
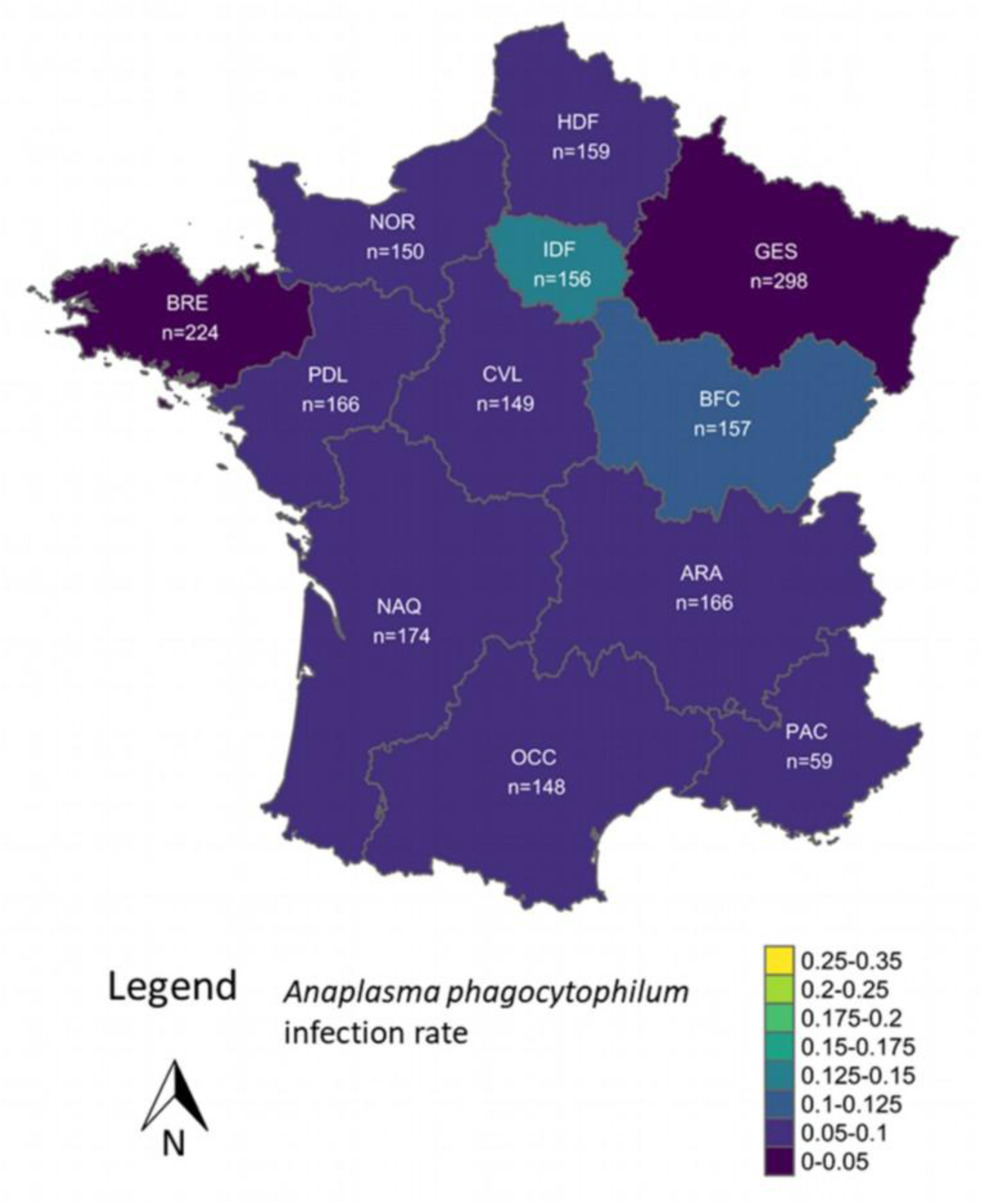
Infection rates of *Anaplasma phagocytophilum* per region in analyzed human-biting ticks. n: number of ticks analyzed in the considered region. Region: 20R: Corse; ARA: Auvergne-Rhône-Alpes; BFC: Bourgogne-Franche-Comté; BRE: Bretagne; CVL: Centre-Val de Loire; GES: Grand-Est; HDF: Hauts-de-France; IDF: Île-de-France; NAQ: Nouvelle-Aquitaine; NOR: Normandie; OCC: Occitanie; PAC: Provence-Alpes-Côte d’Azur; PDL: Pays-de-la-Loire.

### Other microorganisms detected in ticks

Fifty-seven ticks were tested positive for *Ehrlichia* spp., including 50 *Neoerlichia mikurensis,* all detected across different regions of France (Table 4), and exclusively in *I. ricinus*. Among all the other less-frequent pathogens, *Ehrlichia spp.* was the only one detected in the Hauts-de-France region (HDF), with eight positive ticks, including seven *N. mikurensis* (data not shown in Table 4). No *E. canis* was detected.

Thirty-three *Apicomplexa* protozoans were detected across multiple regions, mainly in *I. ricinus* (28 ticks), as well as one *D. marginatus,* one *D. reticulatus* and three ticks of undetermined species (Table 3, Table 4).

Among the 25 *Babesia* spp. detections, five *B. microti* were found in *I. ricinus* from northeastern France: three in GES and two in BFC regions (Table 4). Fourteen *I. ricinus* were infected with *B. venatorum* across France. *Babesia divergens* was detected in five ticks: four *I. ricinus* and one *D. reticulatus*. A single *B. canis*, a known canine pathogen, was detected in an *I. ricinus* tick from the Normandy region (NOR) (Table 3, Table 4).

Two *Theileria* spp. were detected, but the tick species could not be determined.

Two *I. ricinus* and one *D. marginatus* were infected with *Hepatozoon* spp., which are considered as non-pathogenic parasites to humans (Table 3, Table 4). One *Bartonella henselae* was detected in an *I. ricinu*s tick in CVL region (0.7%; 95 CI: 0 - 3.7).

*Babesia ovis, Babesia bovis, Babesia caballi, Mycoplasma* spp.*, Coxiella* spp. were not detected. None of the pathogens listed in Table 4 were detected in ticks collected from regions 20R and PAC that is why these regions were not represented in Table 4.

Fifty-three ticks were infected with a non-pathogenic *Francisella-like* endosymbiont: seven *I. ricinus* with *Francisella*-like type 1; 16 *I. ricinus*, 26 *D. reticulatus*, one *D. marginatus*, one *Hyalomma* spp and two unidentified ticks by *Francisella*-like type 2. They were detected in almost all regions, with infection rates ranging from 0.6% (95 CI: 0 - 3.5) in IDF to 3.6% (95 CI: 1.3 - 7.7) in ARA (Table 4).

**Table 2.** Count and Infection rates of less frequent pathogenic (to humans) and non-pathogenic microorganisms found in human-biting ticks per region. Infection rate are in percentage and 95% confidence interval are between brackets. n: total number of analyzed ticks in each region. Region: ARA: Auvergne-Rhône-Alpes; BFC: Bourgogne-Franche-Comté; BRE: Bretagne; CVL: Centre-Val de Loire; GES: Grand-Est; IDF: Île-de-France; NOR: Normandie; NAQ: Nouvelle-Aquitaine; OCC: Occitanie; PDL: Pays-de-la-Loire. Only regions with less frequent pathogens detected are represented in this table. Pathogens’ higher taxonomical level are in bold letters. Non-pathogenic microorganisms are marked by an asterisk *.

| Pathogens | Regions |  |  |  |  |  |  |  |  |  |
| --- | --- | --- | --- | --- | --- | --- | --- | --- | --- | --- |
|  | ARA<br>n=166 | BFC<br>n=157 | BRE<br>n=224 | CVL<br>n=149 | GES<br>n=298 | IDF<br>n=156 | NOR<br>n=150 | NAQ<br>n=174 | OCC<br>n=148 | PDL<br>n=166 |
| ***Ehrlichia* spp.** | 8; 4.8%<br>(2.1-9.3) | 7; 4.5%<br>(1.8-9) | 2; 0.9%<br>(0.1-3.2) | 7; 4.7%<br>(1.9-9.4) | 8; 2.7%<br>(1.2-5.2) | 1; 0.6%<br>(0-3.5) | 1; 0.7%<br>(0-3.7) | 7; 4%<br>(1.6-8.1) | 6; 4.1%<br>(1.5-8.6) | 2; 1.2%<br>(0.1-4.3) |
| *Neoehrlichia mikurensis* | 8; 4.8%<br>(2.1-9.3) | 7; 4.5%<br>(1.8-9) | 0; 0%<br>(0-1.6) | 6; 4%<br>(1.5-8.6) | 8; 2.7%<br>(1.2-5.2) | 1; 0.6%<br>(0-3.5) | 0; 0%<br>(0-2.4) | 7; 4%<br>(1.6-8.1) | 6; 4.1%<br>(1.5-8.6) | 0; 0%<br>(0-2.2) |
| **Apicomplexa** | 5; 3%<br>(1-6.9) | 4; 2.5%<br>(0.7-6.4) | 5; 2.2%<br>(0.7-5.1) | 2; 1.3%<br>(0.2-4.8) | 4; 1.3%<br>(0.4-3.4) | 3; 1.9%<br>(0.4-5.5) | 3; 2%<br>(0.4-5.7) | 4; 2.3%<br>(0.6-5.8) | 0; 0%<br>(0-2.5) | 3; 1.8%<br>(0.4-5.2) |
| ***Babesia* spp.** | 3; 1.8%<br>(0.4-5.2) | 3; 1.9%<br>(0.4-5.5) | 4; 1.8%<br>(0.5-4.5) | 0; 0%<br>(0-2.4) | 5; 1.7%<br>(0.5-3.9) | 3; 1.9%<br>(0.4-5.5) | 3; 2%<br>(0.4-5.7) | 3; 1.7%<br>(0.4-5) | 0; 0%<br>(0-2.5) | 1; 0.6%<br>(0-3.3) |
| *Babesia microti* | 0; 0%<br>(0-2.2) | 2; 1.3%<br>(0.2-4.5) | 0; 0%<br>(0-1.6) | 0; 0%<br>(0-2.4) | 3; 1%<br>(0.2-2.9) | 0; 0%<br>(0-2.3) | 0; 0%<br>(0-2.4) | 0; 0%<br>(0-2.1) | 0; 0%<br>(0-2.5) | 0; 0%<br>(0-2.2) |
| *Babesia venatorum* | 2; 1.2%<br>(0.1-4.3) | 1; 0.6%<br>(0-3.5) | 4; 1.8%<br>(0.5-4.5) | 0; 0%<br>(0-2.4) | 1; 0.3%<br>(0-1.9) | 2; 1.3%<br>(0.2-4.6) | 1; 0.7%<br>(0-3.7) | 2; 1.1%<br>(0.1-4.1) | 0; 0%<br>(0-2.5) | 1; 0.6%<br>(0-3.3) |
| *Babesia divergens* | 1; 0.6%<br>(0-3.3) | 0; 0%<br>(0-2.3) | 0; 0%<br>(0-1.6) | 0; 0%<br>(0-2.4) | 1; 0.3%<br>(0-1.9) | 1; 0.6%<br>(0-3.5) | 1; 0.7%<br>(0-3.7) | 1; 0.6%<br>(0-3.2) | 0; 0%<br>(0-2.5) | 0; 0%<br>(0-2.2) |
| ***Theileria* spp.\*** | 0; 0%<br>(0-2.2) | 0; 0%<br>(0-2.3) | 1; 0.4%<br>(0-2.5) | 0; 0%<br>(0-2.4) | 0; 0%<br>(0-1.2) | 0; 0%<br>(0-2.3) | 0; 0%<br>(0-2.4) | 1; 0.6%<br>(0-3.2) | 0; 0%<br>(0-2.5) | 0; 0%<br>(0-2.2) |
| ***Hepatozoon* spp.\*** | 1; 0.6%<br>(0-3.3) | 1; 0.6%<br>(0-3.5) | 0; 0%<br>(0-1.6) | 0; 0%<br>(0-2.4) | 0; 0%<br>(0-1.2) | 0; 0%<br>(0-2.3) | 0; 0%<br>(0-2.4) | 0; 0%<br>(0-2.1) | 0; 0%<br>(0-2.5) | 1; 0.6%<br>(0-3.3) |
| ***Francisella*-like endosymbiont\*&** | 6; 3.6%<br>(1.3-7.7) | 4; 2.5%<br>(0.7-6.4) | 8; 3.6%<br>(1.6-6.9) | 3; 2%<br>(0.4-5.8) | 9; 3%<br>(1.4-5.7) | 1; 0.6%<br>(0-3.5) | 3; 2.0%<br>(0.4-5.7) | 5; 2.9%<br>(0.9-6.6) | 2; 1.4%<br>(0.2-4.8) | 5; 3.0%<br>(1-6.9) |

&Total of *Francisella*-like endosymbiont type 1 and 2

### Coinfection

Eighty-one ticks carried two human pathogens (4.0%) and nine carried three pathogens (0.4%) (Tables S4 and S5), representing 4.5% of the total tick sample. Neither developmental stage (Deviance = 3.98, p-value = 0.41) nor species (Deviance = 6.38, p-value = 0.60) significantly improved model fit compared to the null model. We found no statistical evidence that coinfection infection rate differed among stages or species. Coinfections were mostly observed in *I. ricinus* as it was the species most represented in the samples: 80 *I. ricinus* carried two pathogens and eight ones, carried three microorganisms. Two *Dermacentor* were co-infected: one *D. reticulatus* with *B. divergen*s, *A. phagocytophilum*, and *R. raoultii;* and one *D. marginatus* with *A. phagocytophilum* and *R. slovaca*. Coinfection rates with another pathogen among positive ticks were as follows: 26.8% (78/291; 95 CI: 21.8 – 32.3) for *B. burgdorferi* s.l. infected ticks, 54% (27/50; 95 CI: 39.3 – 68.2) for *N. mikurensis* infected ticks, 29.6 % (42/142; 95 CI: 22.2 – 37.8) for *A. phagocytophilum* infected ticks, 25% (30/120; 95 CI: 17.5 – 33.7) for *R. helvetica* infected ticks, and 60% (15/25; 95 CI: 38.7 – 78.9) for *Babesia-*infected ticks. The most frequent association involved *B. afzelii* and *N. mikurensis,* detected together in 20 ticks. Fifteen ticks carried *Babesia* spp. and a second or third microorganism: the involved species were *B. venatorum* (five ticks, including four with *B. burgdorferi* s.l.), *B. microti* (five ticks), *B. divergens* (four ticks), *B. canis* (one tick) (Tables S4 and S5).

## Discussion

This study highlights the value of citizen science to investigate the distribution of biting ticks and their pathogens on a large spatial- scale. By analyzing human-biting ticks using a broad-spectrum targeting method which tests for a range of 43 microorganisms including 20 potential human pathogens, we accessed to the diversity of tick-borne bacterial and protozoan pathogens that humans are exposed to in France through bites of different tick species. Twenty-six different microorganisms were detected in our study, with 18 of them being associated with potential risk for human health, underlining the importance of looking for a large diversity of pathogens. Overall, human pathogens distribution was widespread across the country, and, while there were some spatial variations, no region has been found free of tick-borne pathogens. In our study, some places were studied for the first time thanks to citizen participation, which represents a huge improvement in the knowledge of the distribution of ticks and tick-borne pathogens in France, in addition to what is known in the literature (Perez et al., 2023). Specifically, it confirms the widespread presence of *I. ricinus* throughout the territory, and some of its less-studied pathogens in France, such as *Neorhlichia mikurensis* or *Rickettsia helvetica* (Perez et al., 2023). Some pathogens known to be present in France were not found in our study, like *Francisella tularensis*, *Coxiella burnetti*, *Ehrlichia canis*, *Babesia bovis*. This might be explained by their usually low prevalence, as previously observed in questing ticks (Perez et al., 2023). Additional sampling effort among the biting-tick collection of CiTIQUE should be made to confirm these results; one possibility would be for example to target specific groups like cattle farmers to search for *C. burnetti* and *B. bovis*.

According to our results, the human-biting ticks were mostly *I. ricinus* nymphs, apart from the Mediterranean regions (PAC and COR) where mostly *Dermacentor* and *Rhipicephalus* ticks were found. This corresponds to the expected distribution of these species, especially *I. ricinus* which is less prevalent in southeastern France: the hot and dry climate is less favorable for its development and survival while being better for *Rhipicephalus* ticks (Lebert et al., 2022; Perez et al., 2023; Wongnak et al., 2022). Although we could have expected to have higher number of adult ticks, that are easier to detect by citizens (Eisen and Eisen, 2021), we received *I. ricinus* stages respectively, in proportion similar to other studies on human-biting ticks in other European countries (Mietze et al., 2011; Wilhelmsson et al., 2013; Lernout et al., 2019; Audino et al., 2021; Lista et al., 2022a; Hornok et al., 2025; Koczwarska et al., 2025). This observed distribution of stages is consistent with known biology of *I. ricinus*. Still, if our results do not seem to show a bias toward detecting adult biting tick, it shows a bias towards detecting engorged nymphs compared to engorged adults. Nymphs are smaller and might be more difficult to detect before they reach a sufficient size during blood feeding, while adults are already bigger.

Surprisingly, we found nine nymphs of *D. marginatus* and one nymph of *D. reticulatus*, with their identification confirmed by the microfluidic assay. These nymphs were found during their known seasonal activity (i.e. summer)(Nosek, 1972; Földvári et al., 2016), except for one *D*. *marginatus* nymph that was found in April 2019. In our study, people were mostly bitten in their private yard (except one who was bitten on a trail next to a river). Immature of these tick species are supposed to be endophilic and rarely found questing on vegetation (Rubel et al., 2016a). Previous studies have already found *Dermacentor* nymphs biting humans, and, excepting one *D. reticulatus*, they were always *D. marginatus* (Merino et al., 2005; Andersson et al., 2018; Bakırcı et al., 2019; Lista et al., 2022b; Koczwarska et al., 2023), which is coherent with our results. Compared to *D. reticulatus*, *D. marginatus* is more adapted to warmer and drier climate and can be found in open environments (Rubel et al., 2016b).

Across France, the average infection rate of biting ticks was 27.7%, with infected ticks carrying one to three microorganisms pathogenic to humans. These different pathogens do not have the same implication for human health (from asymptomatic forms to debilitating symptoms), but taken together they represent a global exposure of humans to tick-borne pathogens in France. Still, this infection rate of 27.7 must be interpreted with caution because it does not mean that all people bitten by infected ticks have been infected and developed a disease. Several reasons explain this caution: The method used for pathogen detection is based on DNA amplification and does not distinguish between living and dead microorganisms. Furthermore, as no genotyping analysis was performed, the strains or genotype detected may not have been infectious to humans (Radolf et al., 2021). The method was applied to pathogenic bacteria and protozoa, which are usually transmitted to the host within several hours after starting the blood meal. Pathogen transmission is generally limited if the tick is removed quickly (CDC, 2025). The risk of developing a disease can also be related to human vulnerability (De Keukeleire et al., 2015), for example *B. miyamotoi* is mainly detected in immunocompromised patients (Rubio et al., 2023). We did not find an effect of species or developmental stages on infection probability. The only significant difference was between males and larvae but this result carries substantial uncertainty due to the small number of male ticks and should be interpreted with caution. Moreover, we did not find any effect of engorgement on tick infection rate, suggesting absence of reverse transmission from humans to ticks sampled here. Nonetheless, the percentage of infected biting ticks revealed by this study calls for a reminder of the proper preventive measures against the risk of tick bites and for continuing to raise awareness of this risk among all citizens in France, regardless of where they live.

Unsurprisingly, *Borrelia* was the most frequently detected pathogen, with *B. afzelii* and *B. garinii* being the dominant genospecies. Five regions (ARA, BFC, CVL, GES and NAQ) showed higher *B. burgdorferi* sensu lato infection rates than the others, indicating substantial spatial variation in *Borrelia* distribution across the country. These differences may reflect variation in reservoir host distribution and availability, or potential within-species genetic structuring in *I. ricinus* populations (Gray et al., 2021). This spatial heterogeneity merits further investigation and is presently explored in a subsequent study (Bah et al., 2025).

*Borrelia* was almost absent from samples from the PAC region, due to the fact that the sampling effort was lower and also because the ticks reported belonged to species other than *I. ricinus*. However, an *I. ricinus* tick infected by *Borrelia* was found, confirming the presence of this pathogen in the French Mediterranean region (Sevestre et al., 2021).. Interestingly we found that 7.3 % of the *I. ricinus* larvae were infected by a *B. burgdorferi* s.l. genospecies. Contamination was unlikely, as the same microorganisms were not detected in the samples analyzed just before or after larvae on the PCR microassay and during DNA extraction. . While transovarial transmission of *B. burgdorferi* s.l. was thought to be very unlikely, recent studies have shown that it is possible to find infected questing larvae, and that these infected larvae are even able to infect a new host (Grigoryeva et al., 2024; van Duijvendijk et al., 2016). Larval infection can result from transovarial infection or interrupted feeding, depending on the *Borrelia* genospecies or strain involved. Although unlikely and undocumented, acquisition from humans to larvae cannot be entirely excluded, even if there was no effect of engorgement on tick infection. The different results (Grigoryeva et al., 2024; van Duijvendijk et al., 2016) suggest that larval tick may contribute to Lyme disease transmission risk, which probably require further investigation and evaluation. Prevention messages usually consider larvae as not at risk for Lyme disease transmission, and should be corrected.

Not surprisingly, *Rickettsia* species diversity was highest in the PAC region, where we found a higher diversity of tick vector species. This reinforces results from previous studies (Jumpertz et al., 2023; Parola et al., 2013). *R. helvetica* was the most frequently detected species, consistent with its known association with *I. ricinus*. It has been reported in some clinical cases but its pathogen’s potential is still debated (De Vito et al., 2020). *Rickettsia raoultii* was also detected across all regions of France, albeit at lower rates. This suggests that SENLAT cases can occur nationwide, aligning with the increasing incidence of such cases in recent years in GES region (Kotzyba et al., 2025). Altogether, *Rickettsia*-infected ticks were less frequently detected in the Northwestern regions of France, which likely reflects differences in reservoir host distribution and availability, even if the reservoir hosts of *R. helvetica* are not clearly established (Hornok et al., 2014).

The infection rate of *A. phagocytophilum* was unexpectedly low in the northeastern region (GES), despite it being the region where most diagnoses occur in France (Rigaud et al., 2016). This could be explained by the host specificity of the *A. phagocytophilum* ecotypes, as ecotypes were not characterized and not all strains are associated with human infection but rather primarily infect ruminants (Jaarsma et al., 2019). There is still the possibility of under-reporting of human cases in other regions and/or limited awareness of the disease, as there is no surveillance implemented on this disease. Contamination of our samples appear unlikely, as results were confirmed by two different methods. Conversely, the unexpectedly higher infection rate observed in IDF, may reflect natural fluctuations in prevalence across different years. Some high percentage of *A. phagocytophilum*-infected ticks were indeed found in previous studies in forests of IDF region (Halos et al., 2006; Paul et al., 2016).

Concerning other detected pathogens, *N. mikurensis* was found at low infection rate in almost all regions of France, but nearly none in the northwestern regions. Infection rate is similar to what has been found in questing ticks in a previous study in GES and BRE region (Boyer et al., 2021). As a relatively newly recognized pathogen, it warrants further attention, especially given its non-negligible presence. Its main reservoir hosts are supposed to be small mammals (Burri et al., 2014), which could explain why it is less present in the north-western regions of France. Indeed, our results on pathogens associated to small mammals (*B*. *afzelii*, *R*. *helvetica*, *N*. *mikurensis*), would suggest a lower presence of ticks biting small mammals in north-western regions. This could be due to different small mammal’s communities in this area, with fewer competent host, and/or a higher presence of non-competent host which could be responsible for a dilution effect. Alternatively, it could reflect a difference in the seasonality of *I. ricinus* larval activity and peak activity of small mammals, resulting in larvae feeding preferentially on other type of hosts.

4.5% of ticks were infected by more than one human pathogen. This result might be underestimated if we consider coinfections between *Borrelia* genospecies that are common (Herrmann et al., 2013), and that our method using PCR and sequencing confirmation steps could not determine. However, it actually gives a result that could be representative of coinfection by different potential pathogen species. Indeed, our results are similar to coinfection percentages found in humans (Boyer et al., 2022) but lower than percentages found in some questing ticks that have taken into account coinfections by different *Borrelia* (Lommano et al., 2012; Moutailler et al., 2016). The most common coinfections involved *B. burgdorferi* s.l. and *A. phagocytophilum* or *N. mikurensis*, which were expected given their higher infection rate. Among co-infected ticks, *N. mikurensis*-infected ticks were more frequently co-infected with other pathogens (54%) than ticks infected with other pathogens, suggesting coinfections might be a facilitating factor for *N. mikurensis* to infect ticks. Such a facilitation process has already been suggested for *Babesia* infections (Boyer et al., 2022) which were also mainly found in our study in coinfection with other pathogens (60%). Consequently, when faced with a *N. mikurensis* infection, general practitioners should think of searching for possible coinfections.

From a methodological standpoint, the present approach proved particularly suitable to describe human exposure to *Borrelia* spp. causing Lyme borreliosis, as well as several neglected bacterial and protozoan pathogens transmitted by *I. ricinus* and other tick species. Citizen participation is essential to collect data at a large scale, and their continuing contribution should lead to spatially finer results in coming years that could be useful for other purposes. Indeed, such large-scale dataset represent a valuable resource to support future modeling efforts aiming to better understand the spatiotemporal dynamics of human exposure to tick-borne pathogens. Furthermore, improving sampling representativeness among the CiTIQUE biting tick collection through a more stratified and regionally balanced strategy, would further improve the depth and the robustness of such analyses. This could also be strengthened by improving citizen enrollment in underrepresented regions. Combining citizen-collected human-biting ticks with questing tick surveys, such as with coordinated flagging campaigns, could also provide complementary insights into tick and pathogen distributions. At a very local scale, it might even allow a rigorous comparison between human-biting and questing ticks. This comparison could (i) quantify biases in tick pathogen exposure derived from each sampling method, (ii) compare pathogen diversity between biting and questing ticks, and (iii) test whether carriage of particular pathogens confers ecological or behavioral advantages to the tick.

## Conclusions

This study highlights the power of citizen science for the monitoring- of tick risk and in mapping the diversity and distribution of human tick-borne pathogens across France. While spatial heterogeneity was observed—especially for *Borrelia*, *Rickettsia*, and *Anaplasma* species—no region was free of infected ticks. Specifically, it would be interesting to investigate environmental factors that could explain this spatial heterogeneity. Humans were mostly bitten by *I. ricinus* nymphs, apart from the Mediterranean region. Coinfections, mainly involving *Borrelia* and *Anaplasma* or *Neoehrlichia*, were found in about 5% of ticks, indicating complex pathogen interactions.

However, two emerging public health concerns related to ticks are currently under increased scrutiny in France. The expansion of *Hyalomma* spp., vectors of Crimean-Congo Hemorrhagic Fever virus (CCHFV), highlights the need for targeted surveillance efforts focusing on these species, particularly in regions where they are established or near the current invasion front. Similarly, the ongoing circulation of Tick-Borne Encephalitis virus (TBEV) transmitted by *I. ricinus* raises questions about how the spatial distribution of other tick-borne pathogens may reflect ecological conditions conducive to TBEV transmission. While viral detection remains challenging within the current CiTIQUE framework, due to constraints related to viral genome stability in collected samples, such ecological proxies could help guide future targeted surveillance strategies. These aspects underline the importance of adapting citizen science tools to evolving epidemiological contexts. But it also highlights that future efforts should consider high-risk populations and viral pathogens to refine risk assessments and improve public health responses. In addition, the establishment of a permanent and upkeeping collection of human-biting tick is a distinctive feature of CiTIQUE. By maintaining this resource and making it available to the national and international scientific community, we aim to encourage further research to address new and emerging question in tick-borne disease and public health.

## Data Availability

All data produced in the present study are available upon reasonable request to the authors. Data will be available from the Recherche Data Gouv Repository when accepted.

## Supplementary information

Additional files:

Table S1: Lists of tick species and microorganisms detected by the microfluidic method, their targeted gene and primers

Table S2: List of microorganisms targeted in this study, their known pathogenicity for humans, and their associated competent vector tick species.

Table S3: Number of collected individuals per species and per stages

Table S4: Human-biting ticks carrying two microorganisms.

Table S5: Human-biting ticks carrying three microorganisms

## Data availability

Data will be available from the Recherche Data Gouv Repository here: To be completed when accepted.

## Ethics approval and consent to participate

Approval of the ethics committee was not required since participation in sending ticks was voluntary and the short questionnaire online was anonymous (no identification possible). The participants are given information about the study on the website of the project.

## Declaration of generative AI in scientific writing

During the preparation of this work the authors used ChatGPT and deepL in order to improve English writing. After using this tool/service, the authors reviewed and edited the content as needed and take full responsibility for the content of the published article.

## Acknowledgments

Our thoughts are with Jean-François Cosson (JFC) and Béatrice Palin, who passed away in 2019 and 2022. JFC co-created the CiTIQUE project in 2016 alongside PFK and MVT, and co-designed, with PFK, the foundational structure of the program, including the Signalement TIQUE mobile application and its associated database, which has since become a crucial source of data for research. He also produced and analyzed the initial data that were utilized in this publication. Béatrice Palin was the first “tick librarian” of the program and collaborated with PFK in the design and implementation of the initial procedures for archiving ticks sent in by citizens.

We extend our gratitude to all the citizens and CiTIQUE partners who made this project possible by reporting tick bites and sending the collected ticks to the tick library, as well as to Sandrine Capizzi and the volunteers from CPIE who contributed to raising awareness about the program.

We acknowledge the dedicated work of the tick librarians who have archived all the ticks sent in by citizens (Béatrice Palin, Zoé Gelhaye-Henrion, Sofian Chebli, Antonin Frey, Clémentine Lapie, Tom Beyaert, Aloïs Chevalier, Léane Lémery, Clémentine Frey, Sarah Ost, Eulalie Rathgeber, Sarah Guinet, Sandrine Warion).

We thank Mikaël Carlavan for his assistance in the development of the tick library database alongside JM, as well as Jean-Marc Armand for developing the first version of the Signalement TIQUE application.

## CRediT authorship contribution statement

**Jonas Durand:** Writing – original draft, Writing – review and editing, Conceptualization, Methodology, Formal analysis, Investigation, Data Curation. **Thierno Bah Madiou:** Writing – original draft, Writing – review and editing, Formal analysis, Data curation, Visualization. **Isabelle Lebert:** Writing – original draft, Writing – review and editing, Formal analysis, Data curation. **Clémence Galon:** Writing – review and editing, Methodology, Investigation, Validation. **Irene Carravieri:** Writing – review and editing, Resources. **Sébastien Masseglia:** Writing – review and editing, Validation. **Jean-Marc Armand:** Writing – review and editing, Software, Resources. **Julien Marchand:** Writing – review and editing, Software, Resources. **Cyril Galley:** Writing – review and editing, Resources. **Karine Chalvet-Monfray:** Writing – review and editing, Formal analysis, Supervision. **Muriel Vayssier-Taussat:** Writing – review and editing, Conceptualization, Funding acquisition. **Gwenaël Vourc’h:** Writing – review and editing, Conceptualization. **Annick Brun-Jacob:** Writing – review and editing, Resources, Project administration, Funding acquisition. **Sara Moutailler:** Writing – review and editing, Methodology, Resources, Project administration. **Xavier Bailly:** Writing – review and editing, Formal analysis, Resources, Supervision. **Pascale Frey-Klett:** Writing – review and editing, Conceptualization, Formal analysis, Resources, Supervision, Project administration, Funding acquisition.

