## Supplementary table S1 for "Distribution of tick-borne microorganisms in human-biting ticks in France collected through a Citizen-science program"

**Table S1. Lists of tick species and pathogens, their targeted gene and primers**

| **Organism** | **Targeted gene** | **Primers (F and R - 5'-3') and Probes (P)** | **Product length (bp)** | **Reference** |
| --- | --- | --- | --- | --- |
| *Ixodes ricinus* | *ITS2* | F-CGAAACTCGATGGAGACCTG | 77 | Michelet et al., 2014 |
|  |  | R-ATCTCCAACGCACCGACGT |  |  |
|  |  | P-TTGTGGAAATCCCGTCGCACGTTGAAC |  |  |
| *Dermacentor reticulatus* | *ITS2* | F-AACCCTTTTCCGCTCCGTG | 83 | Michelet et al., 2014 |
|  |  | R-TTTTGCTAGAGCTCGACGTAC |  |  |
|  |  | P-TACGAAGGCAAACAACGCAAACTGCGA |  |  |
| *Dermacentor marginatus* | *ITS2* | F-GCACGTTGCGTTGTTTGCC | 139 | Michelet et al., 2014 |
|  |  | R-CCGCTCCGCGCAAGAATCT |  |  |
|  |  | P-TTCGGAGTACGTCGAGCTCTAGCAGA |  |  |
| Tick species | *16S rRNA* | F-AAATACTCTAGGGATAACAGCGT | 99 | Gondard et al., 2020 |
|  |  | R-TCTTCATCAAACAAGTATCCTAATC |  |  |
|  |  | P-CAACATCGAGGTCGCAAACCATTTTGTCTA |  |  |
| *Escherichia coli* | *eae* | F-CATTGATCAGGATTTTTCTGGTGATA | 102 | Michelet et al., 2014 |
|  |  | R-CTCATGCGGAAATAGCCGTTA |  |  |
|  |  | P-ATAGTCTCGCCAGTATTCGCCACCAATACC |  |  |
| *Borrelia burgdorferi* sensu stricto | *rpoB* | F-GCTTACTCACAAAAGGCGTCTT | 83 | Michelet et al., 2014 |
|  |  | R-GCACATCTCTTACTTCAAATCCT |  |  |
|  |  | P-AATGCTCTTGGACCAGGAGGACTTTCA |  |  |
| *Borrelia garinii* | *rpoB* | F-TGGCCGAACTTACCCACAAAA | 88 | Michelet et al., 2014 |
|  |  | R-ACATCTCTTACTTCAAATCCTGC |  |  |
|  |  | P-TCTATCTCTTGAAAGTCCCCCTGGTCC |  |  |
| *Borrelia afzelii* | *fla* | F-GGAGCAAATCAAGATGAAGCAAT | 116 | Michelet et al., 2014 |
|  |  | R-TGAGCACCCTCTTGAACAGG |  |  |
|  |  | P-TGCAGCCTGAGCAGCTTGAGCTCC |  |  |
| *Borrelia valaisiana* | *ospA* | F-ACTCACAAATGACAGATGCTGAA | 135 | Michelet et al., 2014 |
|  |  | R-GCTTGCTTAAAGTAACAGTACCT |  |  |
|  |  | P-TCCGCCTACAAGATTTCCTGGAAGCTT |  |  |
| *Borrelia lusitaniae* | *rpoB* | F-CGAACTTACTCATAAAAGGCGTC | 87 | Michelet et al., 2014 |
|  |  | R-TGGACGTCTCTTACTTCAAATCC |  |  |
|  |  | P-TTAATGCTCTCGGGCCTGGGGGACT |  |  |
| *Borrelia spielmanii* | *fla* | F-ATCTATTTTCTGGTGAGGGAGC | 71 | Michelet et al., 2014 |
|  |  | R-TCCTTCTTGTTGAGCACCTTC |  |  |
|  |  | P-TTGAACAGGCGCAGTCTGAGCAGCTT |  |  |
| *Borrelia bissetti* | *rpoB* | F-GCAACCAGTCAGCTTTCACAG | 118 | Michelet et al., 2014 |
|  |  | R-CAAATCCTGCCCTATCCCTTG |  |  |
|  |  | P-AAAGTCCTCCCGGCCCAAGAGCATTAA |  |  |
| *Borrelia myamotoi* | *glpQ* | F-CACGACCCAGAAATTGACACA | 94 | Michelet et al., 2014 |
|  |  | R-GTGTGAAGTCAGTGGCGTAAT |  |  |
|  |  | P-TCGTCCGTTTTCTCTAGCTCGATTGGG |  |  |
| *Borrelia* spp. | *23S rRNA* | F-GAGTCTTAAAAGGGCGATTTAGT | 73 | Michelet et al., 2014 |
|  |  | R-CTTCAGCCTGGCCATAAATAG |  |  |
|  |  | P-AGATGTGGTAGACCCGAAGCCGAGT |  |  |
| *Anaplasma marginale* | *msp1* | F-CAGGCTTCAAGCGTACAGTG | 85 | Michelet et al., 2014 |
|  |  | R-GATATCTGTGCCTGGCCTTC |  |  |
|  |  | P-ATGAAAGCCTGGAGATGTTAGACCGAG |  |  |
| *Anaplasma platys* | *groEL* | F-TTCTGCCGATCCTTGAAAACG | 75 | Michelet et al., 2014 |
|  |  | R-CTTCTCCTTCTACATCCTCAG |  |  |
|  |  | P-TTGCTAGATCCGGCAGGCCTCTGC |  |  |
| *Anaplasma phagocytophilum* | *msp2* | F-GCTATGGAAGGCAGTGTTGG | 77 | Michelet et al., 2014 |
|  |  | R-GTCTTGAAGCGCTCGTAACC |  |  |
|  |  | P-AATCTCAAGCTCAACCCTGGCACCAC |  |  |
| *Anaplasma ovis* | *msp4* | F-TCATTCGACATGCGTGAGTCA | 92 | Michelet et al., 2014 |
|  |  | R-TTTGCTGGCGCACTCACATC |  |  |
|  |  | P-AGCAGAGAGACCTCGTATGTTAGAGGC |  |  |
| *Anaplasma centrale* | *groEL* | F-AGCTGCCCTGCTATACACG | 79 | Michelet et al., 2014 |
|  |  | R-GATGTTGATGCCCAATTGCTC |  |  |
|  |  | P-CTTGCATCTCTAGACGAGGTAAAGGGG |  |  |
| *Anaplasma bovis* | *groEL* | F-GGGAGATAGTACACATCCTTG | 73 | Sprong et al. 2019 |
|  |  | R-CTGATAGCTACAGTTAAGCCC |  |  |
|  |  | P-AGGTGCTGTTGGATGTACTGCTGGACC |  |  |
| *Anaplasma* spp. | *16S rRNA* | F-CTTAGGGTTGTAAAACTCTTTCAG | 160 | Gondard et al., 2020 |
|  |  | R-CTTTAACTTACCAAACCGCCTAC |  |  |
|  |  | P-ATGCCCTTTACGCCCAATAATTCCGAACA |  |  |
| *Ehrlichia canis* | *gltA* | F-GACCAAGCAGTTGATAAAGATGG | 136 | Gondard et al., 2020 |
|  |  | R-CACTATAAGACAATCCATGATTAGG |  |  |
|  |  | P-ATTAAAACATCCTAAGATAGCAGTGGCTAAGG |  |  |
| *Ehrlichia* spp. | *16S rRNA* | F-GCAACGCGAAAAACCTTACCA | 98 | Gondard et al., 2020 |
|  |  | R-AGCCATGCAGCACCTGTGT |  |  |
|  |  | P-AAGGTCCAGCCAAACTGACTCTTCCG |  |  |
| *Neoehrlichia mikurensis* | *groEL* | F-AGAGACATCATTCGCATTTTGGA | 96 | Michelet et al., 2014 |
|  |  | R-TTCCGGTGTACCATAAGGCTT |  |  |
|  |  | P-AGATGCTGTTGGATGTACTGCTGGACC |  |  |
| *Rickettsia conorii* | *23S-5S ITS* | F-CTCACAAAGTTATCAGGTTAAATAG | 118 | Michelet et al., 2014 |
|  |  | R-CGATACTCAGCAAAATAATTCTCG |  |  |
|  |  | P-CTGGATATCGTGGCAGGGCTACAGTAT |  |  |
| *Rickettsia slovaca* | *23S-5S ITS* | F-GTATCTACTCACAAAGTTATCAGG | 138 | Michelet et al., 2014 |
|  |  | R-CTTAACTTTTACTACAATACTCAGC |  |  |
|  |  | P-TAATTTTCGCTGGATATCGTGGCAGGG |  |  |
| *Rickettsia massiliae* | *23S-5S ITS* | F-GTTATTGCATCACTAATGTTATACTG | 128 | Michelet et al., 2014 |
|  |  | R-GTTAATGTTGTTGCACGACTCAA |  |  |
|  |  | P-TAGCCCCGCCACGATATCTAGCAAAAA |  |  |
| *Rickettsia helvetica* | *23S-5S ITS* | F-AGAACCGTAGCGTACACTTAG | 79 | Michelet et al., 2014 |
|  |  | R-GAAAACCCTACTTCTAGGGGT |  |  |
|  |  | P-TACGTGAGGATTTGAGTACCGGATCGA |  |  |
| *Rickettsia aeshlimannii* | *23S-5S ITS* | F-CTCACAAAGTTATCAGGTTAAATAG | 134 | Sprong et al., 2018 |
|  |  | R-CTTAACTTTTACTACGATACTTAGCA |  |  |
|  |  | P-TAATTTTTGCTGGATATCGTGGCGGGG |  |  |
| *Rickettsia felis* | *orfB* | F-ACCCTTTTCGTAACGCTTTGC | 163 | Gondard et al., 2020 |
|  |  | R-TATACTTAATGCTGGGCTAAACC |  |  |
|  |  | P-AGGGAAACCTGGACTCCATATTCAAAAGAG |  |  |
| *Rickettsia* spp. | *gltA* | F-GTCGCAAATGTTCACGGTACTT | 145 | Michelet et al., 2014 |
|  |  | R-TCTTCGTGCATTTCTTTCCATTG |  |  |
|  |  | P-TGCAATAGCAAGAACCGTAGGCTGGATG |  |  |
| *Bartonella henselae* | *pap31* | F-CCGCTGATCGCATTATGCCT | 107 | Michelet et al., 2014 |
|  |  | R-AGCGATTTCTGCATCATCTGCT |  |  |
|  |  | P-ATGTTGCTGGTGGTGTTTCCTATGCAC |  |  |
| *Bartonella* spp. | *ssrA* | F-CGTTATCGGGCTAAATGAGTAG | 118 | Gondard et al., 2020 |
|  |  | R-ACCCCGCTTAAACCTGCGA |  |  |
|  |  | P-TTGCAAATGACAACTATGCGGAAGCACGTC |  |  |
| *Francisella tularensis* | *tul4* | F-ACCCACAAGGAAGTGTAAGATTA | 76 | Michelet et al., 2014 |
|  |  | R-GTAATTGGGAAGCTTGTATCATG |  |  |
|  |  | P-AATGGCAGGCTCCAGAAGGTTCTAAGT |  |  |
| *Francisella-*like endosymbiont | *fop4* | F-GGCAAATCTAGCAGGTCAAGC | 91 | Michelet et al., 2014 |
|  |  | R-CAACACTTGCTTGAACATTTCTAG |  |  |
|  |  | P-AACAGGTGCTTGGGATGTGGGTGGTG |  |  |
| *Coxiella burnetii* | *IS1111* | F-TGGAGGAGCGAACCATTGGT | 86 | Michelet et al., 2014 |
|  |  | R-CATACGGTTTGACGTGCTGC |  |  |
|  |  | P-ATCGGACGTTTATGGGGATGGGTATCC |  |  |
| *Coxiella-*like endosymbiont | *idc* | F-AGGCCCGTCCGTTATTTTACG | 74 | Michelet et al., 2014 |
|  |  | R-CGGAAAATCACCATATTCACCTT |  |  |
|  |  | P-TTCAGGCGTTTTGACCGGGCTTGGC |  |  |
| Apicomplexa | *18S* | F-TGAACGAGGAATGCCTAGTATG | 104 | Gondard et al., 2020 |
|  |  | R-CACCGGATCACTCGATCGG |  |  |
|  |  | P-TAGGAGCGACGGGCGGTGTGTAC |  |  |
| *Babesia microti* | *CCTeta* | F-ACAATGGATTTTCCCCAGCAAAA | 145 | Michelet et al., 2014 |
|  |  | R-GCGACATTTCGGCAACTTATATA |  |  |
|  |  | P-TACTCTGGTGCAATGAGCGTATGGGTA |  |  |
| *Babesia canis* (3 subspecies) | *18 rRNA* | F-TGGCCGTTCTTAGTTGGTGG | 104 | Michelet et al., 2014 |
|  |  | R-AGAAGCAACCGGAAACTCAAATA |  |  |
|  |  | P-ACCGGCACTAGTTAGCAGGTTAAGGTC |  |  |
| *Babesia ovis* | *18S rRNA* | F-TCTGTGATGCCCTTAGATGTC | 92 | Michelet et al., 2014 |
|  |  | R-GCTGGTTACCCGCGCCTT |  |  |
|  |  | P-TCGGAGCGGGGTCAACTCGATGCAT |  |  |
| *Babesia bovis* | *CCTeta* | F-GCCAAGTAGTGGTAGACTGTA | 100 | Michelet et al., 2014 |
|  |  | R-GCTCCGTCATTGGTTATGGTA |  |  |
|  |  | P-TAAAGACAACACTGGGTCCGCGTGG |  |  |
| *Babesia caballi* | *Rap1* | F-GTTGTTCGGCTGGGGCATC | 94 | Michelet et al., 2014 |
|  |  | R-CAGGCGACTGACGCTGTGT |  |  |
|  |  | P-TCTGTCCCGATGTCAAGGGGCAGGT |  |  |
| *Babesia venatorum* (sp. EU1) | *18S rRNA* | F-GCGCGCTACACTGATGCATT | 91 | Michelet et al., 2014 |
|  |  | R-CAAAAATCAATCCCCGTCACG |  |  |
|  |  | P-CATCGAGTTTAATCCTGTCCCGAAAGG |  |  |
| *Babesia divergens* | *hsp70* | CTCATTGGTGACGCCGCTA | 83 | Michelet et al., 2014 |
|  |  | R-CTCCTCCCGATAAGCCTCTT |  |  |
|  |  | P-AGAACCAGGAGGCCCGTAACCCAGA |  |  |
| *Theileria* spp. | *18S rRNA* | F-GTCAGTTTTTACGACTCCTTCAG | 213 | Melis et al., 2024 |
|  |  | R-CCAAAGAATCAAGAAAGAGCTATC |  |  |
|  |  | P-AATCTGTCAATCCTTCCTTTGTCTGGACC |  |  |
| *Hepatozoon* spp. | *18S rRNA* | F-ATTGGCTTACCGTGGCAGTG | 175 | Gondard et al., 2020 |
|  |  | R-AAAGCATTTTAACTGCCTTGTATTG |  |  |
|  |  | P-ACGGTTAACGGGGGATTAGGGTTCGAT |  |  |
| *Mycoplasma* spp | 16S | F-GTGACGGCTAACTATGTGCC | 77 | Banović et al., 2024 |
|  |  | R-GCTTTACGCCCAATAATTCCG |  |  |
|  |  | P-AGCAGCTGCGGTAATACATAGGTCGC |  |  |
