## Supplementary table S2 for "Distribution of tick-borne microorganisms in human-biting ticks in France collected through a Citizen-science program"

**Table S2. Human pathogenicity and suspected competent tick vector species of the different microorganisms targeted in the study. List of competent tick vector species is based only on the different tick species found in the study and is not valid for other tick species.**

| **Microorganism** | **Human pathogenicity** | **Suspected competent tick vector species** | **Bibliography** |
| --- | --- | --- | --- |
| ***Borrelia* spp** |  |  |  |
| *Borrelia burgdorferi* sensu lato |  | *Ixodes ricinus***, Ixodes hexagonus** | Human pathogenicity: (Stanek and Reiter, 2011; Strnad et al., 2023)  Vector competence: (Eisen, 2020) |
| *Borrelia burgdorferi* sensu stricto | Yes | *Ixodes ricinus***, Ixodes hexagonus** | Human pathogenicity: (Stanek and Reiter, 2011; Strnad et al., 2023)  Vector competence: (Eisen, 2020) |
| *Borrelia garinii* | Yes | *Ixodes ricinus***, Ixodes hexagonus* | Human pathogenicity: (Stanek and Reiter, 2011; Strnad et al., 2023)  Vector competence: (Eisen, 2020) |
| *Borrelia afzelii* | Yes | *Ixodes ricinus***, Ixodes hexagonus* | Human pathogenicity: (Stanek and Reiter, 2011; Strnad et al., 2023)  Vector competence: (Eisen, 2020) |
| *Borrelia valaisiana* | No | *Ixodes ricinus, Ixodes hexagonus* | Human pathogenicity: (Margos et al., 2017; Stanek and Reiter, 2011; Strnad et al., 2023)  Vector competence: (Eisen, 2020) |
| *Borrelia lusitaniae* | Suspected | *Ixodes ricinus, Ixodes hexagonus* | Human pathogenicity: (Stanek and Reiter, 2011; Strnad et al., 2023)  Vector competence: (Eisen, 2020) |
| *Borrelia spielmanii* | Suspected | *Ixodes ricinus, Ixodes hexagonus* | Human pathogenicity: (Stanek and Reiter, 2011; Strnad et al., 2023)  Vector competence: (Eisen, 2020) |
| *Borrelia bissettii* | Suspected | *Ixodes ricinus, Ixodes hexagonus* | Human pathogenicity: (Stanek and Reiter, 2011; Strnad et al., 2023)  Vector competence: (Eisen, 2020) |
| *Borrelia miyamotoi* | Yes | *Ixodes ricinus* | Human pathogenicity and vector competence: (Boulanger et al., 2025) |
| ***Anaplasma* spp** |  |  |  |
| *Anaplasma marginale* | No | *Rhipicephalus sanguineus, Rhipicephalus bursa, Hyalomma spp.* | Human pathogenicity and vector competence: (Rymaszewska and Grenda, 2008) |
| *Anaplasma platys* | No | *Rhipicephalus sanguineus, Hyalomma spp.* | Human pathogenicity and vector competence: (Rymaszewska and Grenda, 2008) |
| *Anaplasma phagocytophilum* | Yes | *Ixodes ricinus* | Human pathogenicity and vector competence: (Rymaszewska and Grenda, 2008) |
| *Anaplasma ovis* | No | *Rhipicephalus sanguineus, Rhipicephalus bursa* | Human pathogenicity and vector competence: (Rymaszewska and Grenda, 2008) |
| *Anaplasma centrale* | No | *Ixodes spp.*, *Haemaphysalis spp.* | Human pathogenicity and vector competence: (Rymaszewska and Grenda, 2008) |
| *Anaplasma bovis* | No | *Haemaphysalis* *spp*., Dermacentor spp., Ixodes spp., Hyalomma spp. | Human pathogenicity and vector competence: (Rymaszewska and Grenda, 2008) |
| **Ehrlichia spp** |  |  |  |
| *Neoehrlichia mikurensis* | Yes | *Ixodes ricinus* | Human pathogenicity and vector competence: (Boulanger et al., 2025) |
| **Rickettsia spp** |  |  |  |
| *Rickettsia conorii* | Yes | *Rhipicephalus sanguineus* | Human pathogenicity and vector competence: (Parola et al., 2013) |
| *Rickettsia slovaca* | Yes | *Dermacentor marginatus*, *Dermacentor reticulatus* | Human pathogenicity and vector competence: (Parola et al., 2013) |
| *Rickettsia massiliae* | Yes | *Rhipicephalus sanguineus*, *Ixodes ricinus* | Human pathogenicity and vector competence: (Parola et al., 2013) |
| *Rickettsia helvetica* | Suspected | *Ixodes ricinus* | Human pathogenicity and vector competence: (Parola et al., 2013) |
| *Rickettsia aeschlimannii* | Yes | *Hyalomma marginatum, Hyalomma rufipes* | Human pathogenicity and vector competence: (Parola et al., 2013) |
| *Rickettsia felis* | No | *Ixodes* *ricinus*, *Ixodes* *hexagonus* | Human pathogenicity and vector competence: (Parola et al., 2013) |
| *Rickettsia raoultii* | Yes | *Dermacentor marginatus*, *Dermacentor reticulatus* | Human pathogenicity and vector competence: (Parola et al., 2013) |
| *Rickettsia monacensis* | Suspected | *Ixodes ricinus* | Human pathogenicity and vector competence: (Parola et al., 2013) |
| ***Bartonella spp*** |  |  |  |
| *Bartonella henselae* | Yes | Not vectorized by ticks to humans. | Human pathogenicity and vector competence: (Torrejón et al., 2022) |
| ***Francisella tularensis****^&^* | **Yes** | *Ixodes ricinus*, *Dermacentor marginatus*, *Dermacentor reticulatus* | Human pathogenicity and vector competence: (Boulanger et al., 2025) |
| ***Coxiella burnetii****^&^* | **Yes** | *Ixodes ricinus*, *Dermacentor marginatus*, *Dermacentor reticulatus*, *Hyalomma spp.*, *Haemaphysalis spp*. | Human pathogenicity and vector competence: (Boulanger et al., 2025) |
| ***Babesia spp*** |  |  |  |
| *Babesia microti* | Yes | *Ixodes ricinus* | Human pathogenicity: (Bajer et al., 2022)  Vector competence: (Bajer and Dwużnik-Szarek, 2021) |
| *Babesia canis* | No | *Dermacentor reticulatus* | Human pathogenicity: (Bajer et al., 2022)  Vector competence: (Bajer and Dwużnik-Szarek, 2021) |
| *Babesia ovis* | No | *Rhipicephalus bursa* | Human pathogenicity: (Bajer et al., 2022)  Vector competence: (Bajer and Dwużnik-Szarek, 2021) |
| *Babesia bovis* | No | *Dermacentor marginatus* | Human pathogenicity: (Bajer et al., 2022)  Vector competence: (Bajer and Dwużnik-Szarek, 2021) |
| *Babesia caballi* | No | *Rhipicephalus bursa* | Human pathogenicity: (Bajer et al., 2022)  Vector competence: (Bajer and Dwużnik-Szarek, 2021) |
| *Babesia venatorum* | Yes | *Ixodes ricinus* | Human pathogenicity: (Bajer et al., 2022)  Vector competence: (Bajer and Dwużnik-Szarek, 2021) |
| *Babesia divergens* | Yes | *Ixodes ricinus* | Human pathogenicity: (Bajer et al., 2022)  Vector competence: (Bajer and Dwużnik-Szarek, 2021) |
| ***Theileria spp*** | **No** | *Dermacentor reticulatus*, *Rhpicephalus spp.* | Human pathogenicity: (Bajer et al., 2022)  Vector competence: (Bajer and Dwużnik-Szarek, 2021) |
| ***Mycoplasma spp*** | **No** | *Rhipicephalus spp.*, *Hyalomma spp.* | Human pathogenicity and vector competence: (Migliore et al., 2024; Shi et al., 2025; Soliman et al., 2025) |
| ***Hepatozoon spp*** | **No** | *Ixodes spp., Hyalomma spp., Haemaphysalis spp., Rhipicephalus spp.* | Human pathogenicity and vector competence: (Uiterwijk et al., 2023) |

^&^The main mode of transmission of this pathogen to humans is not through ticks.

* strong evidence for vector competence based on a combination of in vitro studies on tick to host and host to tick transmission.
