## Supplementary tables S3, S4 and S5 for "Distribution of tick-borne microorganisms in human-biting ticks in France collected through a Citizen-science program"

**Table S3 : Number of collected individuals per species and per stages**

**M: male adult tick; F: Female adult tick; N: nymph; L: larva**

|  | **Developmental stages** | | | | |
| --- | --- | --- | --- | --- | --- |
| **Species** | **M** | **F** | **N** | **L** | **Undetermined** |
| *Dermacentor marginatus* | 4 | 15 | 9 | 0 | 5 |
| *Dermacentor reticulatus* | 3 | 13 | 1 | 0 | 10 |
| *Haemaphysalis punctata* | 0 | 1 | 0 | 0 | 0 |
| *Hyalomma spp* | 0 | 1 | 0 | 0 | 0 |
| *Ixodes hexagonus* | 0 | 1 | 4 | 0 | 0 |
| *Ixodes ricinus* | 3 | 218 | 1,324 | 110 | 236 |
| *Rhipicephalus bursa* | 0 | 1 | 0 | 0 | 0 |
| *Rhipicephalus sanguineus* | 3 | 6 | 1 | 0 | 0 |
| *Unidentified* | 2 | 16 | 7 | 2 | 13 |

**Table S4: Human-biting ticks carrying two pathogens.**  **The pathogens with an asterisk were found in *Dermacentor marginatus***

| **Microorganisms_1** | **Microorganisms_2** | **Co_Occurrences** |
| --- | --- | --- |
| *Borrelia burgdorferi sensu lato* | *Anaplasma phagocytophilum* | 18 |
| *Borrelia burgdorferi sensu lato* | *Neoehrlichia mikurensis* | 18 |
| *Borrelia burgdorferi sensu lato* | *Rickettsia helvetica* | 12 |
| *Borrelia burgdorferi sensu lato* | *Borrelia miyamotoi* | 4 |
| *Borrelia burgdorferi sensu lato* | *Babesia venatorum* | 4 |
| *Borrelia burgdorferi sensu lato* | *Babesia microti* | 3 |
| *Borrelia burgdorferi sensu lato* | *Bartonella henselae* | 1 |
| *Borrelia miyamotoi* | *Anaplasma phagocytophilum* | 2 |
| *Borrelia miyamotoi* | *Neoehrlichia mikurensis* | 1 |
| *Anaplasma phagocytophilum* | *Rickettsia helvetica* | 4 |
| *Anaplasma phagocytophilum* | *Neoehrlichia mikurensis* | 3 |
| *Anaplasma phagocytophilum* | *Babesia divergens* | 2 |
| *Anaplasma phagocytophilum* | *Rickettsia raoultii* | 2 |
| *Anaplasma phagocytophilum* | *Rickettsia monacensis* | 1 |
| *Anaplasma phagocytophilum** | *Rickettsia slovaca** | 1* |
| *Neoehrlichia mikurensis* | *Rickettsia helvetica* | 3 |
| *Rickettsia helvetica* | *Babesia venatorum* | 1 |
| *Rickettsia helvetica* | *Babesia divergens* | 1 |

**Table S5: Human-biting ticks carrying three pathogens. Pathogens with an asterisk were found in *Dermacentor reticulatus*.**

| **Microorganisms_1** | **Microorganisms_2** | **Microorganisms_3** | **Co_Occurrences** |
| --- | --- | --- | --- |
| *Borrelia burgdorferi sensu lato* | *Anaplasma phagocytophilum* | *Rickettsia helvetica* | 3 |
| *Borrelia burgdorferi sensu lato* | *Babesia microti* | *Rickettsia helvetica* | 2 |
| *Borrelia burgdorferi sensu lato* | *Neoehrlichia mikurensis* | *Rickettsia helvetica* | 2 |
| *Borrelia burgdorferi sensu lato* | *Anaplasma phagocytophilum* | *Borrelia miyamotoi* | 1 |
| *Anaplasma phagocytophilum** | *Babesia divergens** | *Rickettsia raoultii** | 1 |
